# Evaluation of imputation performance of multiple reference panels in a Pakistani population

**DOI:** 10.1101/2023.12.22.23300448

**Authors:** Jiayi Xu, Dongjing Liu, Arsalan Hassan, Giulio Genovese, Alanna C. Cote, Brian Fennessy, Esther Cheng, Alexander W. Charney, James A. Knowles, Muhammad Ayub, Roseann E. Peterson, Tim B. Bigdeli, Laura M. Huckins

## Abstract

Genotype imputation is crucial for GWAS, but reference panels and existing benchmarking studies prioritize European individuals. Consequently, it is unclear which publicly available reference panel should be used for Pakistani individuals, and whether ancestry composition or sample size of the panel matters more for imputation accuracy. Our study compared different reference panels to impute genotype data in 1814 Pakistani individuals, finding the best performance balancing accuracy and coverage with meta-imputation with TOPMed and the expanded 1000 Genomes (ex1KG) reference. Imputation accuracy of ex1KG outperformed TOPMed despite its 30-fold smaller sample size, supporting efforts to create future panels with diverse populations.

## Background

Genotype imputation allows easy, accurate, and significant increases in the number of variants available for genome wide association studies (GWAS), facilitating data harmonization and meta-analyses across cohorts and genotyping platforms, as well as statistical fine-mapping (1).

Currently, >85% of GWAS comprise individuals of European ancestry (EUR) (2), and available reference panels are predominantly European. Inclusion of non-EUR populations is critical to increase equity in genetic research (3) and advance understanding of genetic architecture of disease. Individuals of South Asian (SAS) ancestry, especially those in Pakistan, are severely under-represented in genetic studies, with few published GWAS (4). While imputation accuracy has been evaluated in an Indian population (5), no study has yet assessed imputation accuracy in Pakistani individuals based on reference panels (i.e., TOPMed (6), high-coverage expanded 1000 Genomes (ex1KG) (7), low-coverage 1000 Genomes (1KG) (8), GenomeAsia (9)). These panels include a small proportion of Pakistani samples, suggesting limited utility to studies of Pakistani populations and as-yet-determined imputation accuracy. Although TOPMed is by far the largest imputation panel (N = 97,256), only 0.1% are Pakistani individuals (n=139) (6,10). The ex1KG and GenomeAsia-Pilot panels have 146 and 113 Pakistani samples with a total sample size of 3,202 and 1,739, respectively (7,9).

Here, we compare accuracy of imputation panels in a cohort of 1814 Pakistani individuals. We include comparison of true R^2^ (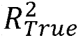 through leveraging targeted sequencing data) and estimated R^2^ 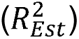. We assess 5 imputation panels, including TOPMed, ex1KG, 1000G GRCh38 (1KG38), 1000G GRCh37 SAS (1KG37-SAS), GenomeAsia-Pilot, as well as a meta-imputation approach combining TOPMed and ex1KG (meta). In this study, we for the first time examine which of the publicly available panels yields the best performance for Pakistani populations, whether merging existing panels further improves imputation accuracy, and whether ancestry or sample size is a more critical determinant of imputation accuracy.

## Results and Discussion

We initially evaluated imputation accuracy among common variants (minor allele frequency (MAF) ≥ 1% in the Pakistani individuals). To evaluate the imputation accuracy against true genotypes, 1748 Pakistani individuals in this study (96%) also have sequenced genotype data via targeted sequencing. When comparing the imputed genotypes to targeted sequencing data, broadly, there was no difference in imputation accuracy for common variants across the six panels except for MAF bins of (0.01, 0.015] and (0.4, 0.5] (Figure 1a, average 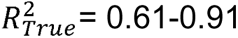 across MAF bins of common variants). Genome-wide, including variants for which we do not have matched targeted sequencing data, ex1KG and meta-imputation had the highest imputation accuracy 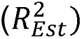 for common variants (mean 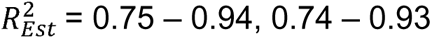 respectively), while GenomeAsia-Pilot and 1KG37-SAS panels had the lowest imputation accuracy (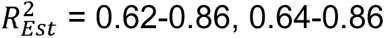 respectively, Figures 2a). ex1KG also had the highest imputation accuracy measured via empirical R^2^ 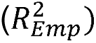 for single nucleotide polymorphisms (SNPs) that were both imputed and genotyped on the Illumina Infinium Global Screening Array (GSA) (Fig. S9).

**Figure 1.**
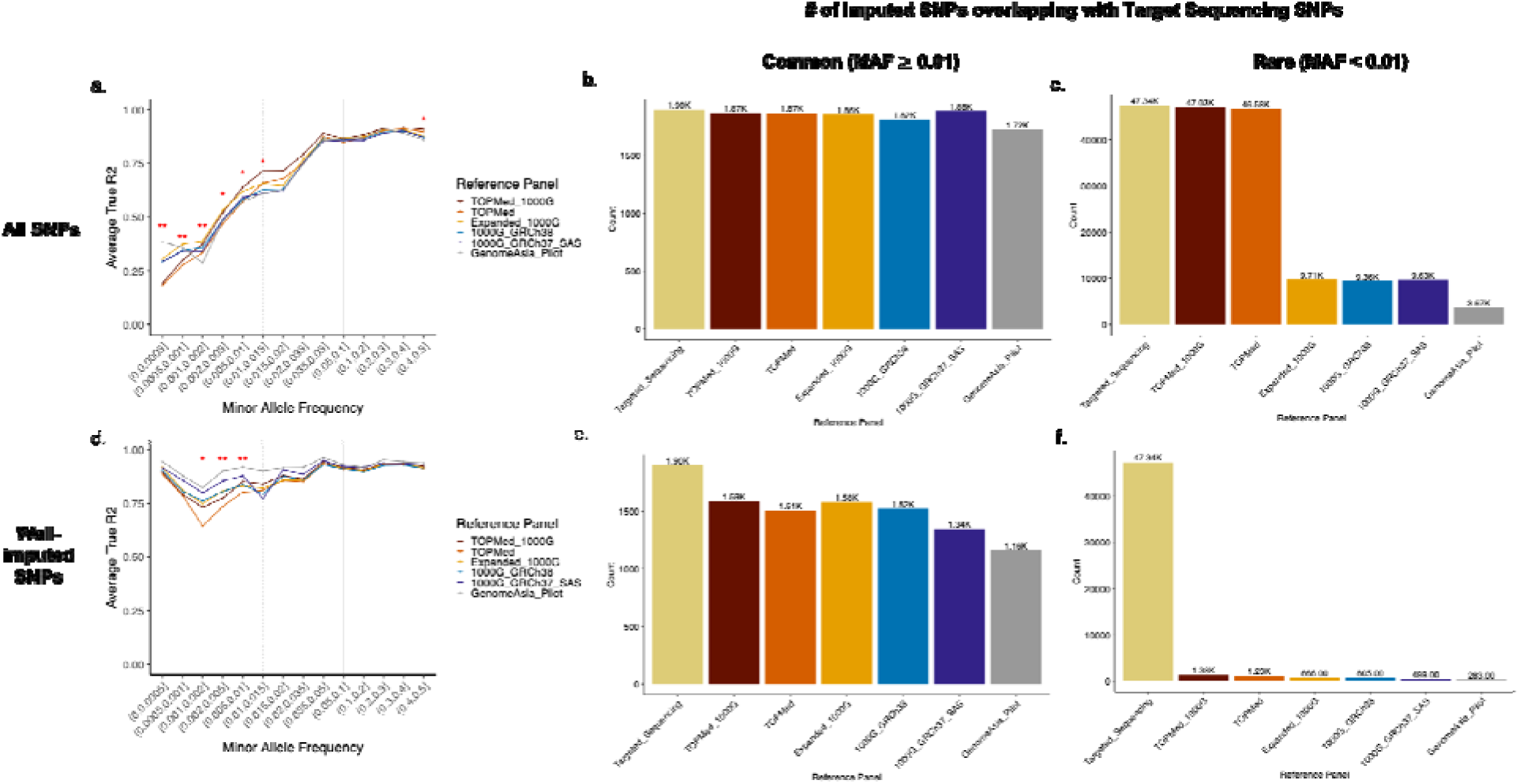
Imputation accuracy and coverage compared against targeted sequencing in the Pakistani individuals. a) Imputation accuracy of different imputation panels across different MAF bins, measured by, for variants that were also included in the targeted sequencing (i.e., 49234 exonic variants on 161 genes). is the squared correlation between imputed genotype dosage and sequenced genotype, with 1 being a perfect match. Significance: * indicates a nominally significant difference in the mean across different imputation panels (Kruskal-Wallis rank sum test P < 0.05) at a specific MAF bin; ** denotes a Bonferroni-corrected significant difference (P < 0.003, correcting for the number of pairwise panels tested). The horizontal grey lines indicate of 0.6 (dashed line, indicating fairly high imputation quality) and 0.8 (solid line, indicating high imputation quality). The vertical grey lines indicate MAF > 1% (dashed line) and > 5% (solid line); b) The number of common variants imputed by different panels compared to targeted sequencing; c) The number of rare variants imputed by different panels compared to targeted sequencing; d) Imputation accuracy of different imputation panels, measured by, for variants with an of 0.8 or greater. SNPs with high 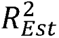 also have high 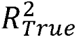 values (average > 0.65), suggesting the appropriateness of using 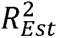 as a proxy for 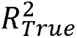 when there is no sequencing data available as the gold standard reference; e) The number of well-imputed common variants by different imputation panels compared against targeted sequencing; f) The number of well-imputed rare variants (defined as 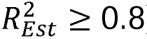) by different imputation panels compared against targeted sequencing. Abbreviation: 1000G, 1000 Genomes; MAF, minor allele frequency; SAS, South Asian ancestry; SNP, single nucleotide polymorphism.

**Figure 2.**
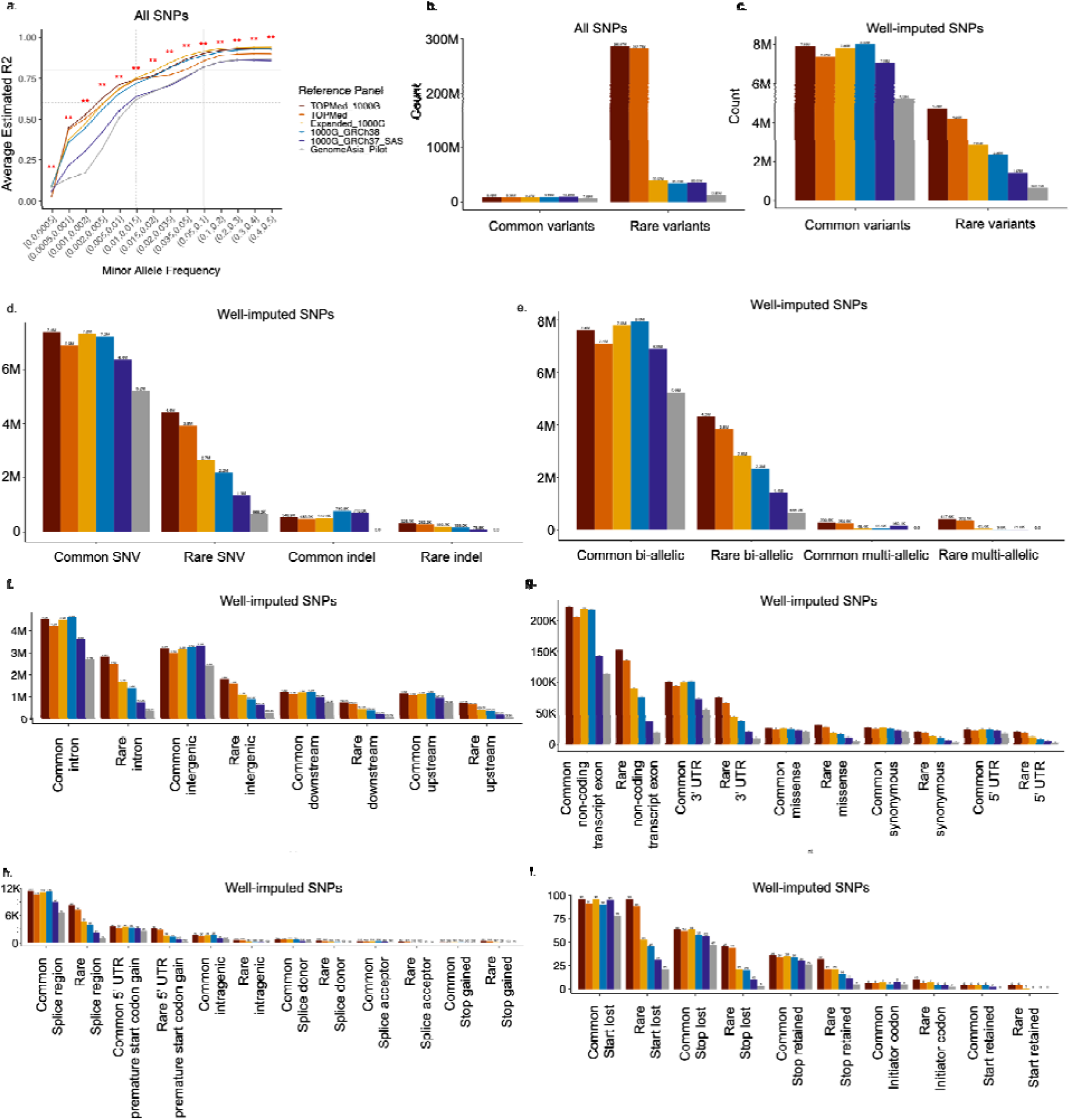
Genome-wide imputation accuracy and coverage in the Pakistani individuals. a) Imputation accuracy of different imputation panels measured by, which is the squared correlation between imputed genotype dosage and true genotype based on posterior allele probabilities. A value closer to 1 indicates higher imputation accuracy. Significance: * indicates a nominally significant difference in the mean across different imputation panels (Kruskal-Wallis rank sum test P < 0.05) at a specific MAF bin; ** denotes a Bonferroni-corrected significant difference (P < 0.003, correcting for the number of pairwise panels tested). The horizontal grey lines indicate 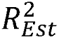 of 0.6 (dashed line, indicating fairly high imputation quality) and 0.8 (solid line, indicating high imputation quality). The vertical grey lines indicate MAF > 1% (dashed line) and > 5% (solid line); b) The number of imputed SNPs genome-wide by common/rare variants; c) The number of well-imputed SNPs (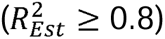) genome-wide by common/rare variants; d) The number of well-imputed SNPs genome-wide by common/rare and SNV/indel status; e) The number of well-imputed SNPs genome-wide by common/rare and bi-allelic/multi-allelic status; f-i) The number of well-imputed SNPs genome-wide by common/rare status and SNP types via functional annotation; f) includes 4 SNP types with the highest SNP counts (i.e., intron, intergenic, downstream, upstream); g) includes 5 SNP types with following highest SNP counts (i.e., non-coding transcript exon, 3’ UTR, missense, synonymous, 5’ UTR); h) includes 6 SNP types with low SNP counts (i.e., splice region, 5’ UTR premature start codon gain, intragenic, splice donor, splice acceptor, stop gained); i) includes 5 SNP types with ultra-low SNP counts (i.e., start lost, stop lost, stop retained, initiator codon, start retained). Abbreviation: 1000G, 1000 Genomes; indel, insertion-deletion; MAF, minor allele frequency; SAS, South Asian ancestry; SNP, single nucleotide polymorphism; SNV, single nucleotide variant; UTR, untranslated region.

Next, we evaluated which imputation panel offered the best coverage for imputation of common variants, defined as percentage of targeted sequencing SNPs that were also imputed by a specific imputation panel. 90.9% - 99.4% of common variants called in our targeted sequencing data were included in at least one imputation panel (Figure 1b), of which 61.3% (GenomeAsia-Pilot) to 83.8% (meta-imputation) were well-imputed 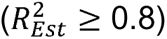 (Figure 1e). In particular, among the 1896 common variants through targeted sequencing, 1589 were well-imputed by meta-imputation, followed by ex1KG with 1578 SNPs. On a genome-wide scale, imputation with 1KG38 resulted in the most well-imputed SNPs (8.02M), followed by meta-imputation (7.93M) and ex1KG (7.85M); for average 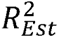, ex1KG is the best, followed by meta-imputation and then 1KG38. Imputation with GenomeAsia-Pilot led to the fewest imputed and well-imputed SNPs (Figures 1b, 1e, 2b, 2c).

Overall, for common variants, balancing genomic coverage and imputation accuracy, meta-imputation had the best performance, followed by ex1KG which had the highest imputation accuracy but slightly fewer well-imputed variants (7.85M vs 7.93M). However, meta-imputation is computationally intensive and expensive in terms of both time and data storage, taking ∼70 hours (and 328G disk space) compared to ∼2 hours (and 87G) for ex1KG for the 1814 Pakistani individuals in this study (Table S3).

We found that imputation quality was low for rarer variants (MAF < 1%) irrespective of the imputation reference panel used. However, for lower frequency variants with MAF between 0.5% and 1%, average 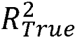 and 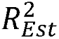 were acceptable (> 0.6) for imputed SNPs using meta-imputation and/or ex1KG (Figs. 1a, 2a). When comparing against sequenced data, meta-imputation and ex1KG had the highest mean 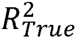 for SNPs with MAF 0.1% - 1% (*p*_meta vs ex1KG_ > 0.35, Figure 1a). On a genome-wide level, meta-imputation had the highest mean 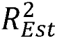 for SNPs with MAF 0.05% −1%. For the lowest MAF category (MAF ≤ 0.05%), findings were inconsistent: GenomeAsia-Pilot had the highest mean 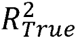, while 1KG38 had the highest mean 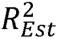, but both had poor imputation quality on an absolute scale (0.38 for GenomeAsia-Pilot 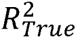, 0.092 for 1KG38 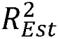). It is likely that the mean 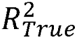 and 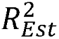 for meta-imputation was diluted at the ultra-rare end due to its substantial number of poorly imputed rare variants 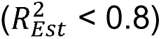 compared to other imputation panels (Fig. S10).

Imputation coverage was also low for rare variants. Although meta-imputation and TOPMed imputation yielded 98.4%-99.4% of targeted sequencing rare variants (Figure 1c), this number drastically dropped when including only well-imputed SNPs (2.6%-2.9% of targeted sequencing variants, Figure 1f). Therefore, sequencing remains the best approach to retrieve high-quality genotypes for rare variants. Nonetheless, we still obtained a large absolute number of high-quality genotypes for rare variants through imputation (e.g., 4.8M genome-wide rare variants via meta-imputation for Pakistani individuals). Across imputation panels, meta-imputation had the highest count of both imputed and well-imputed rare variants (286.9M, 4.8M respectively), followed by TOPMed (282.8M, 4.2M). 1KG37-SAS and GenomeAsia-Pilot had the fewest well-imputed variants (1.43M and 0.7M; Figs. 2b, 2c).

Overall, for rare variants, meta-imputation yielded the highest imputation quality and generated the highest count of well-imputed SNPs, followed by imputation with TOPMed. However, meta-imputation required ∼70 hours of computational time, compared to ∼5 hours for TOPMed for the 1814 Pakistani individuals (Table S3). Further, meta-imputation required ∼328G of disk space, compared to ∼150G using TOPMed. We do not recommend the current GenomeAsia-Pilot and 1KG37-SAS datasets for imputation of rare variants given the relatively low counts of well-imputed rare variants obtained.

Next, we tested whether SNPs with greater MAF differences between Pakistani and EUR populations had significant differences in imputation accuracy (measured by 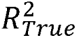), since most reference panels included in this study are primarily of European origin (with the exception of GenomeAsia-Pilot). In fact, deviation from EUR MAF did not have a negative impact on SNP imputation quality (Figs. S11-13), across both common and rare variants. Since all the imputation panels included in this study have 0.1% - 6.5% sequenced individuals from Pakistan (n ranges from 113 to 146 for Pakistani individuals, and *n* ranges from 489 to 724 for SAS individuals in general; Table S1), this suggests that imputation quality is unlikely to impaired by a large proportion of EUR ancestry in reference panels as long as there is some inclusion of individuals from the targeted population (11).

For comparison of coverage across different SNP categories, we mainly focused on well-imputed SNP counts since these are the ones typically included in GWAS (Figs. 2d-2i, S14-S15). For common variants, meta-imputation imputed the most SNVs (7.4M) whereas 1KG38 imputed the most indels (780.8K) (Fig. 2d). Meta-imputation also produced the highest SNP count in most of the SNP categories for common variants, including SNVs (7.4M, Fig. 2d), missense variants, non-coding transcript exonic variants, variants in splice region, and start/stop variants (Fig. 2g-2i), whereas 1KG38 produced the highest SNP count for indels (780.8K; Fig. 2d), intronic variants, and upstream and downstream gene variants (Fig. 2f). For rare variants, meta-imputation produced the highest SNP counts across all categories, followed by TOPMed. Regardless of common or rare variants, imputation using GenomeAsia-Pilot produced the least number of well-imputed SNPs across different SNP types, with no imputed indel or multi-allelic variants. Notably, although far fewer high-quality rare variants are available through imputation panels compared to sequencing (Fig. 1f), on a genome-wide scale, current imputation panels still provide a substantial number of well-imputed rare variants across different SNP categories (Fig. 2d-2i) for further investigation. For example, meta-imputation generated 4.75 M well-imputed rare variants, including 29933 well-imputed rare missense variants (compared to 24616 well-imputed common missense variants). Given high rate of consanguineous marriage increases the probability of homozygous rare variants, genetic studies of rare variants in the Pakistani population as well as other South Asian populations could provide valuable insights into genetic etiology of various diseases for future therapeutic development (5).

Together, our results imply that meta-imputation is currently the best choice for imputation of both common and rare SNPs in Pakistani individuals, taking into account both imputation accuracy and variant coverage. Other panels may also be appropriate; for example, ex1KG has high imputation accuracy for common variants, and lower computational demands, while TOPMed has high coverage of well-imputed rare variants. To address the question of whether sample size or ancestry matching matters more for imputation in the Pakistani population, our findings suggest that ancestry matters more for genetic imputation of common variants, as ex1KG outperformed TOPMed for the number of well-imputed common SNPs (7.85M vs 7.37M, Figs. 1e, 2c) with a smaller total sample size (N_ex1KG_= 3202 vs N_TOPMed_ = 97256) but a larger Pakistani-specific sample size (N_ex1KG_= 146 vs N_TOPMed_ = 139). For rare variants, while whole genome sequencing remains the gold standard to obtain a large number of rare variants, the absolute number of well-imputed SNPs genome-wide by imputation is still massive (>4M for TOPMed and meta-imputation, Fig. 2c) and more cost-friendly compared to whole genome sequencing (on a relative scale of $100 vs $1000 per sample). While sample size is important to increase the likelihood of finding certain rare variants in the population, ancestry is also important for Pakistani-specific rare variants given the high rate of consanguineous marriages (12). As GWAS diversity initiatives gain momentum in the recent years, it is crucial to explore the impact of reference panel ancestry and sample size on imputation. Our study highlighted the significance of these factors in the context of genetic research diversification using the Pakistani population as a case study.

Further, although the GenomeAsia-Pilot had lower performance amongst all the imputation panels tested in this study, a future release of its whole reference panel of 100K (9), assuming 6.5% of Pakistani individuals, would provide the largest Pakistani-specific imputation panel (estimated N = 6500 *versus* ex1KG which has 146 Pakistani individuals, Table S1). In addition, with the option of meta-imputation, imputation power could be further improved by meta-imputing the future release of GenomeAsia with other large panels (e.g., TOPMed). Another alternative approach to improve imputation accuracy while controlling the sequencing cost in the future is to sequence a subset of population-specific individuals and meta-impute with other large reference panels (13).

## Conclusions

In this study, we evaluated imputation performance of various imputation panels in the Pakistani population for the first time. Overall, we found meta-imputation to have the highest genome-wide imputation accuracy for rare variants whereas ex1KG had the highest genome-wide imputation accuracy for common variants. Balancing imputation quality with genomic coverage, our study shows that meta-imputation of TOPMed and ex1KG is the best for imputation of both rare and common variants in Pakistani individuals when the computational resources are not limited. Otherwise, we recommend TOPMed for rare variant imputation and ex1KG for common variant imputation. Further, this study suggests that SNPs with MAF deviated from EUR MAF did not have reduced imputation quality in Pakistani individuals. Increased European sample sizes in imputation reference panels seem unlikely to increase imputation accuracy in non-Europeans; we found the ex1KG panel, with fewer overall individuals but more Pakistani individuals, outperformed the overall larger TOPMed panel for common variants. Taken together, our study supports the importance of including more diverse populations into the current repertoire of human genome reference panels for future genetic research.

## Methods

### Study Subjects

The Pakistani individuals included in this study were collected as pilot in preparation for the Genetics of Schizophrenia in Pakistan (GEN-SCRIP) Study. The samples were collected by Lahore Institute of Research and Development and University of Peshawar. The diagnoses were made based on clinician’s interview according to the Diagnostic and Statistical Manual of Mental Disorders IV. The study was approved by the Institutional Review Board (IRB) committee at University of Peshawar in Pakistan, the IRB of Lahore Institute of Research and Development and IRB of University of Health Sciences Lahore.

### Genotyping and Quality Control

The Infinium^TM^ Global Screening Array-24 v3.0 BeadChip (Illumina Inc., California, USA) was used for genotyping of the Pakistani individuals. Genotyping was performed at the Genomic Core Facility of Icahn School of Medicine at Mount Sinai. The MoChA pipeline was used on Google Cloud to convert the Illumina genotype intensity files (.idat) to VCF files via Illumina GenomeStudio (14,15) (Fig. S1, Supplemental methods). Genetic variants with GenTrain score < 0.4 and cluster separation score < 0.3 were excluded to remove variants with poor genotype cluster separation (Figs. S2, S3). Quality control (QC) was then performed using PLINK (16). At the SNP level, we removed SNPs with low call rate (<0.95), duplicated SNPs with a lower call rate, and those with low minor allele frequency (MAF < 0.001) (Figs. S2, S3). Additionally, given that Pakistan has a tradition of consanguineous marriages which could result in a high level of autozygosity in the population, we also excluded variants that deviate from Hardy-Weinberg Equilibrium (HWE, p<1×10^-6^) only in individuals with low autozygosity (Fig. S3) (4). Low autozygosity is defined as an individual’s consanguinity coefficient F_ROH_ less than 0.5% (4) (F_ROH_ = ∑ L_ROH_ /L_genome_, where L_ROH_ is the sum of the length of all runs of homozygosity (ROH) detected in a subject, and L_genome_ is the total length of the human genome, estimated to be 2772.7 megabases (17)). ROH was called via the PLINK command ---homozyg on a pruned set of common autosomal SNPs with pair-wise correlation <0.8 (--indep-pairwise 50 10 0.8, --maf 0.01). At the individual level, we removed samples with high missingness (>5%) and high heterozygosity (F_het_ < 0.23 or > 0.33, Figs. S2, S4), individuals with a lower call rate for duplicated sample pairs, related individuals (3^rd^ degree or closer with a kinship coefficient > 0.088; those with the highest call rate were retained), those with ambiguous sex (0.2 < F estimate < 0.8), and any samples indicative of swap/mismatch issues (Figs. S2, S4). After the thorough QC, a total of 520,234 variants were retained for 1814 Pakistani samples for phasing (Fig. S2). In addition, the principal component analysis (PCA) was constructed, and all study subjects were confirmed to have South Asian ancestry (Fig. S5).

### Phasing and Imputation

Phasing was performed based on the 1000 Genome Project Phase 3 GRCh38 reference panel (8) using SHAPEIT4 via the MoChA pipeline on Google Cloud (14,15) to generate phased vcf files (Fig. S1, Supplemental methods). The MoChA pipeline has its default filtering steps to exclude SNPs with more than 3% missingness and SNPs with excess heterozygosity (i.e., p<1×10^-6^ in a HWE test) before the phasing process. Next, we uploaded the sorted chromosome-separated phased VCF files to the TOPMed and Michigan imputation servers (18) for imputation via Minimac4 version 1.7.3. The imputation reference panels evaluated in this study include TOPMed (GRCh38) (6), ex1KG (GRCh38) (7), 1KG38 (GRCh38) (8), 1KG37-SAS (GRCh37) (8) and GenomeAsia-Pilot (GRCh37) (9). The sample size, in particular SAS and Pakistani sample size, in each imputation panel as well as the mean read depth are included in Table S1.

In addition, we performed meta-imputation using both TOPMed and ex1KG imputed results through the MetaMinimac2 tool, which was designed to improve imputation performance by statistically combined imputed results from different reference panels without the need to physically merging the imputation panels together (19). Since meta-imputation can only run on imputed results of the same genome build, we selected TOPMed (GRCh38) and ex1KG (GRCh38) for meta-imputation, given that TOPMed is currently the largest imputation reference panel whereas ex1KG has the largest sample size of Pakistani individuals (Table S1). An overview of the study design for this paper is outlined in Fig. S1.

### Targeted Sequencing

Besides genotyping on SNP arrays, targeted sequencing of 49234 exonic variants on 161 genes on 22 autosomal chromosomes was also performed in these Pakistani individuals, using the Ion Torrent platform at Sema4, Inc. (Mount Sinai Genomics Inc., Connecticut, USA) with an average sequencing depth of 224x (20). The Ion AmpliSeq technology was used to create the sequencing library, in which the amplicons for the 161 genes were designed using Ion AmpliSeq Designer version 6.13. Individual genotype called from the targeted sequencing was considered as the true genotype to assess the accuracy of imputed genotype using different imputation reference panels. The detailed QC of the targeted sequencing data is described elsewhere (20).

### Imputation accuracy evaluation

#### True R-square

The true R^2^ 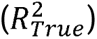 is the squared correlation between the imputed genotype dosage and the sequenced genotype. It is considered as the most robust measure to assess the imputation accuracy given it is based on comparison against directly measured genotype from targeted sequencing with rich read depth (mean: 224x), compared to the mean read depth used for included imputation reference panels (7.4x to 38.2x, Table S1). Theoretically, 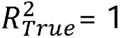 indicates that the imputed genotypes are the same as the sequenced genotypes, and therefore a 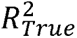 value closer to 1 suggests a higher imputation accuracy. 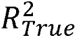 was calculated for each individual SNP by the aggRSquare tool (19) and averaged across the default MAF bins in the aggRSquare tool, including MAF cutoffs of 0.0005, 0.001, 0.002, 0.005, 0.010, 0.015, 0.020, 0.035, 0.05, 0.1, 0.2, 0.3, 0.4, and 0.5. Given we only have targeted sequencing data for exonic variants of 161 genes, the 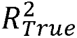 can only be evaluated on a subset of genome-wide SNPs on the 22 autosomal chromosomes (n=49234).

#### Estimated R-square

Without sequenced genotypes as a gold standard reference, R^2^ may be estimated 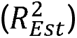 based on their posterior allele probabilities for each imputed SNP on a genome-wide scale (Table S2). Calculation of 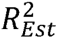 is an embedded function in the Minimac tool used by the online imputation server (11) as well as in the MetaMinimac2 tool for meta-imputation (19). When Hardy Weinberg equilibrium holds (11), 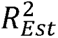 is equivalent to the INFO score produced by the IMPUTE software (21), another imputation quality score frequently used for SNP QC for GWAS.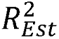 may be less accurate than 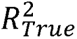 when there are insufficient samples for imputation (11), and/or when the SNPs are rare (correlation with 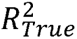 dropped to 0.72-0.77 for rare variants across imputation panels, Fig. S6). Well-imputed SNPs in this study are defined as 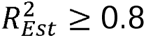.

To test whether 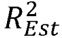 is an appropriate proxy for 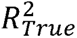 in our study, we calculated imputation accuracy for SNPs with high 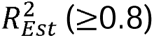, a filter often applied in GWAS, and observed a corresponding improvement in 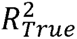 across MAF, with most mean 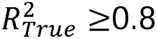 across different MAF bins (Figure 1d; correlation of 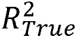 and 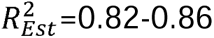, Fig. S6), confirming that 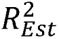 can be used as a proxy for 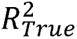 when there is no sequencing data available to serve as reference (Fig. S7).

#### Empirical R-Square

Empirical R^2^ 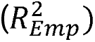 is the squared correlation between the imputed genotype dosage and the directly measured genotype on the GSA array. Imputed SNP dosages of these genotyped loci were calculated by masking the measured genotype as if they were unknown and then imputing these SNPs. Therefore, 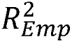 can be calculated only for SNPs directly genotyped on the GSA array (i.e., nearly 500K SNPs) (Table S2). The 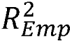 calculation is included in Minimac, but not in the MetaMinimac2 tool for meta-imputation (19). 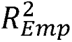 was not strongly correlated with 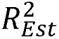 (*r* = 0.43-0.54 across imputation panels, Figs. S8-S9) and therefore it is a less accurate measure for imputation accuracy, compared to 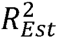, which has a high correlation with 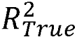. Table S4 explains the difference between the three types of R^2^.

### SNP type

In this study, we defined rare variants as MAF < 0.01, and common if MAF ≥ 0.01. The distinction between single nucleotide variant (SNV) and insertion-deletions (indel) was based on the number of nucleotides at one locus (i.e., if both reference and alternate alleles are single nucleotides, then it was classified as a SNV; if any of the alleles had > 1 nucleotide, it was classified as an indel). We used SnpEff to functionally annotate all the imputed variants (22). For imputed data using the GenomeAsia-Pilot and 1KG37-SAS panels, the GRCh37.75 version of the pre-built human database was used for annotation, whereas for imputed data using other imputation panels, the GRCh38.99 version was used.

### Statistical analyses

To compare imputation accuracy across different imputation panels at each MAF bin, we used the Kruskal-Wallis rank sum test for significance testing, which is a non-parametric test of one-way ANOVA when ANOVA assumptions (i.e., homogeneity of variance and normality) are not met. Further, to test for pairwise significance, we applied a pairwise t test without assuming equal variance in the two groups, setting two-sided nominal significance at 0.05 for all statistical analyses, while Bonferroni correction was applied to correct for multiple testing to reduce false positive findings (e.g., Bonferroni-corrected significance set at P_Bonferroni_ = 0.05/15 = 0.003 for pairwise comparison across 6 imputation panels). To evaluate whether SNPs with allele frequency (AF) deviated from EUR AF would have lower imputation quality, we performed linear regressions between true R^2^ and the absolute difference of AF using SAS AF based on targeted sequencing and 1000G-based EUR AF. All analyses were conducted in the R statistical software (version 4.2.0, Vienna, Austria).

## Data Availability

Datasets used in this study are not publicly available because it contains private patient data in Pakistan, but they are available from the corresponding author on request. The source code for the analyses is available and deposited in GitHub (https://github.com/xuj18/).

## Declarations

## Ethics approval and consent to participate

Interviewers at the hospitals/enrollment centers from Pakistan collected informed consent from the participants. The study was approved by the Ethics committee at University of Peshawar, University of Health Sciences, Lahore and Lahore Institute of Research and Development in Pakistan.

## Author contributions

J.X. analyzed the data and drafted the manuscript for this study. J.X., R.E.P., T.B.B., and L.M.H. conceptualized and designed the study. J.X. and L.M.H. revised the manuscript. A.H., J.A.K., and M.A. initiated the GEN-SCRIP study and collected data including blood samples from the study participants in Pakistan. D.L., B.F., E.C., and A.W.C. contributed to the production of both genotyping and targeted sequencing data for these Pakistani individuals. D.L. provided targeted sequencing data with quality control for the analysis in this manuscript. G.G. provided consultation on the use of MoChA pipeline. A.C.C. contributed to figure production in this manuscript. All authors read and approved the final manuscript for submission.

## Acknowledgement

This work was supported in part through the computational resources and staff expertise provided by Scientific Computing at the Icahn School of Medicine at Mount Sinai. Research reported in this paper was supported by the Office of Research Infrastructure of the National Institutes of Health under award number S10OD018522 and S10OD026880. The content is solely the responsibility of the authors and does not necessarily represent the official views of the National Institutes of Health. We would also like to thank Dr. Ketian Yu in the Department of Biostatistics at University of Michigan-Ann Arbor for her technical assistance with the MetaMinimac2 tool and the aggRSquare tool.

