## Supplementary Information for "Evaluation of imputation performance of multiple reference panels in a Pakistani population"

**Table of Content**

|  | Page |
| --- | --- |
| Supplemental Methods | 3 |
| Supplemental Tables | 5 |
| Supplemental Figures | 11 |
| Supplemental References | 30 |

**Supplemental Methods**

MoChA Pipeline

The parameters below were used to convert the Illumina idat files to vcf files using the MoChA pipeline [1] on the Google Cloud.

{

"mocha.sample_set_id": "pakistan_gsa",

"mocha.mode": "idat",

"mocha.target": "vcf",

"mocha.realign": true,

"mocha.max_win_size_cm": 300.0,

"mocha.overlap_size_cm": 5.0,

"mocha.ref_name": "GRCh38",

"mocha.ref_path": "gs://bigdeli-working/pakistan/GRCh38",

"mocha.manifest_path": "gs://bigdeli-working/pakistan/manifests",

"mocha.batch_tsv_file": "gs://bigdeli-working/pakistan/samples/pakistan_gsa.batch.tsv",

"mocha.sample_tsv_file": "gs://bigdeli-working/pakistan/samples/pakistan_gsa.sample.tsv",

"mocha.data_path": "gs://bigdeli-working/pakistan/idats"

}

After the manual quality control (QC) on the genotype data, the script below was used for phasing using the MoChA pipeline.

{

"mocha.sample_set_id": "pakistan_gsa_match",

"mocha.mode": "vcf",

"mocha.max_win_size_cm": 50.0,

"mocha.overlap_size_cm": 5.0,

"mocha.ref_name": "GRCh38",

"mocha.ref_path": "gs://bigdeli-working/pakistan/GRCh38",

"mocha.batch_tsv_file": "gs://bigdeli-working/pakistan/samples/gsa_1814/pakistan_gsa.batch.tsv",

"mocha.sample_tsv_file": "gs://bigdeli-working/pakistan/cromwell/outputs/gsa_2433_vcf/pakistan_gsa.sample.tsv",

"mocha.data_path": "gs://bigdeli-working/pakistan/cromwell/outputs/gsa_2433_vcf/",

"mocha.extra_xcl_vcf_file": "gs://bigdeli-working/pakistan/samples/gsa_1814/pakistan_gsa_removed_SNP.recode.vcf",

"mocha.duplicate_samples_file": "gs://bigdeli-working/pakistan/samples/gsa_1814/pakistan_gsa_removed_619_samples.tsv"

}

| **Reference Panel** | **Mean read depth** | **Total sample size** | **AFR (%)** | **AMR (%)** | **EAS (%)** | **EUR (%)** | **SAS (%)** | **SAS: Pakistani (%)** | **Others (%)** |
| --- | --- | --- | --- | --- | --- | --- | --- | --- | --- |
| TOPMed [2] | 38.2x | 97256 | 24267  (25.0%) | 17085 (17.6%) | 1184  (1.2%) | 47159 (48.5%) | 644  (0.7%) | 139  (0.1%) | 6917  (7.1%) |
| Expanded 1000 Genomes [3] | 30x | 3202 | 893  (27.9%) | 490  (15.3%) | 585  (18.3%) | 633  (19.8%) | 601  (18.8%) | 146  (4.6%) | NA |
| 1000 Genomes [4] | 7.4x | 2504 | 661  (26.4%) | 347  (13.9%) | 504  (20.1%) | 503  (20.1%) | 489  (19.5%) | 113  (4.5%) | NA |
| GenomeAsia Pilot [5] | 36x | 1739 | 104  (6.0%) | 26  (1.5%) | 697 (40.1%) | 114  (6.6%) | 724  (41.6%) | 113  (6.5%) | 74  (4.3%) |

**Table S1. Mean read depth, total sample size and breakdown of sample size per ancestry in each imputation reference panel.** The proportion per ancestry group is included in parenthesis for each reference panel, summing up to 100%. The number of Pakistani individuals is also included as part of the number of SAS individuals. For the TOPMed reference panel [2], 6917 individuals are included in the other category as they cannot be assigned to any of these five super-populations. For the GenomeAsia Pilot panel [5], the other category refers to 74 Oceania individuals. Abbreviation: AFR, African ancestry; AMR, admixed American ancestry; EAS, East Asian ancestry; EUR, European ancestry; NA, not applicable; SAS: South Asian ancestry.

| **Reference Panel** | Human genome reference build | # imputed SNPs | # well-imputed SNPs with $R_{Est}^{2}$ $\geq$ 0.8 | # genotyped SNPs with empirical R^2^ |
| --- | --- | --- | --- | --- |
| Meta-imputation (TOPMed + Expanded 1000 Genomes) | GRCh38 | 296353412 | 12679760 | NA |
| TOPMed | GRCh38 | 292136462 | 11573395 | 487217 |
| Expanded 1000 Genomes | GRCh38 | 48891239 | 10705550 | 484357 |
| 1000 Genomes | GRCh38 | 45207412 | 10365883 | 487835 |
| 1000 Genomes SAS | GRCh37 | 47099551 | 8503043 | 491089 |
| GenomeAsia Pilot | GRCh37 | 21494648 | 5896149 | 461435 |

**Table S2. Number of imputed SNPs, well-imputed SNPs, and genotyped SNPs with empirical R^2^ values based on each imputation reference panel.** The well-imputed SNPs are defined as those with estimated R^2^ ($R_{Est}^{2}$) greater or equal to 0.8. The $R_{Est}^{2}$ is an estimation of imputation accuracy based on posterior allele probabilities. The $R_{Emp}^{2}$ is the squared correlation between the imputed genotype dosage and the directly measured genotype on the GSA array. The number of imputed SNPs also includes the number of genotyped SNPs imputed through masking. Abbreviation: GSA, the Illumina Infinium^TM^ Global Screening Array; NA, not applicable; SAS, South Asian ancestry; SNP, single nucleotide polymorphism.

| **Reference Panel** | **Computation time for imputation** |
| --- | --- |
| Meta-imputation (TOPMed + Expanded 1000 Genomes) | 62 h 17 min 52 sec |
| TOPMed | 5 h 34 min 23 sec |
| Expanded 1000 Genomes | 2 h 19 min 5 sec |
| 1000 Genomes | 3 h 56 min 48 sec |
| 1000 Genomes SAS | 3 h 47 min 19 sec |
| GenomeAsia Pilot | 2 h 30 min 43 sec |

**Table S3. Computational time for imputation**. The meta-imputation was performed on Minerva, the high-performance computing cluster at Icahn School of Medicine at Mount Sinai. The meta-imputation took up 6 GB memory (maximum), 7 threads (maximum), and 5 processes (maximum). All the remaining imputation were performed using the online imputation servers (Michigan imputation server: https://imputationserver.sph.umich.edu/index.html; TOPMed imputation server: https://imputation.biodatacatalyst.nhlbi.nih.gov). Note that besides the imputation time per se, there may be time waiting in the queue for the imputation depending on how many jobs other users have submitted on the online imputation server, and therefore the queue time may vary from instant imputation to several days. Abbreviation: SAS, South Asian ancestry.

| **Type of R^2^** | **Definition** | **Pros** | **Cons** |
| --- | --- | --- | --- |
| True R^2^  ($R_{True}^{2}$) | Squared correlation between imputed genotype and sequenced genotype | Most Accurate | Not genome-wide (49235 SNPs) |
| Estimated R^2^ ($R_{Est}^{2}$) | Estimated squared correlation between imputed genotype and true genotype based on posterior allele probabilities [6] | Genome-wide | No sequence data available as gold standard comparison. |
| Empirical R^2^  ($R_{emp}^{2}$) | Squared correlation between imputed genotype and directly measured genotype on the GSA arrays | Theoretically, this should be an accurate measure; but this is not supported by our data. | Not genome-wide (approximately 480000 SNPs); Not applicable to meta-imputation; the least accurate. |

**Table S4. Differences between three types of R^2^ used in the study to examine imputation accuracy.** Abbreviation: GSA, the Illumina Infinium^TM^ Global Screening Array; SNP, single nucleotide polymorphism.

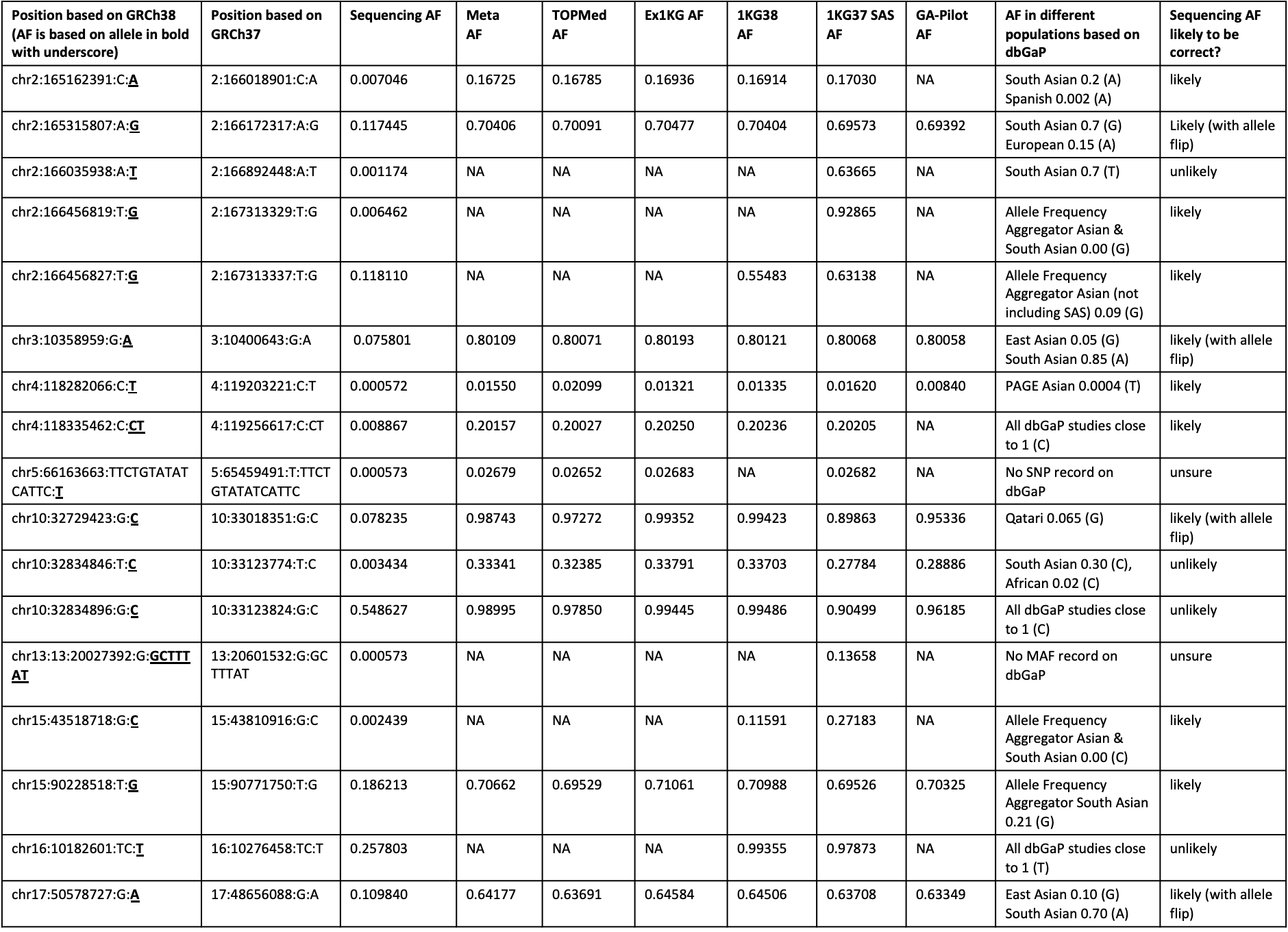

**Table S5. Outlier SNPs with large allele frequency difference between sequenced genotypes and imputed genotypes (n=17).** These SNPs are also visually plotted in Figs. S11-S13. Allele frequency (AF) information in different populations were searched in the dbGaP database (<https://www.ncbi.nlm.nih.gov/gap/>). Based on allele frequency observed in different populations, speculation was made regarding whether the sequencing AF could be correct (i.e., imputed dosage is incorrect for a certain SNP). Overall, sequenced genotypes are likely to be correct for most of these outlier SNPs. Abbreviation: AF, allele frequency; dbGaP, the database of Genotypes and Phenotypes; ex1KG: expanded 1000 Genomes; GA, GenomeAsia; MAF, minor allele frequency; meta, meta-imputation; NA, not applicable (i.e., not imputed by the imputation panel); PAGE, the Population Architecture using Genomics and Epidemiology study; SAS, South Asian ancestry; SNP, single nucleotide polymorphism; 1KG38: 1000 Genomes with GRCh38 genome build; 1KG37: 1000 Genomes with GRCh37 genome build.

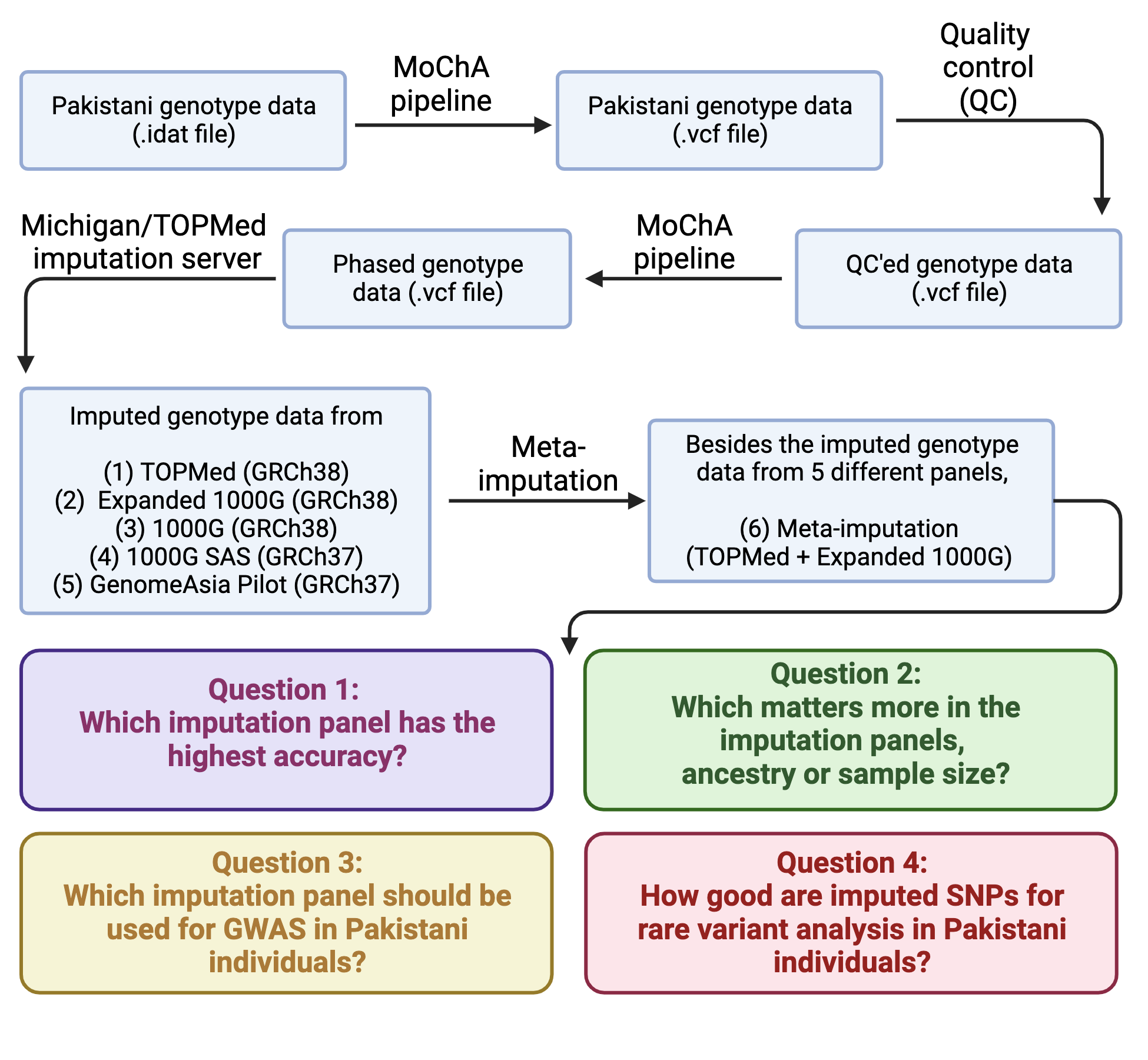
**Figure S1. Study overview flowchart.** Details on how to run the MoChA pipeline on the Google Cloud can be found here: <https://github.com/freeseek/mochawdl>. Abbreviation: 1000G, 1000 Genomes; GWAS, genome-wide association study; SAS, South Asian ancestry; SNP, single nucleotide polymorphism.

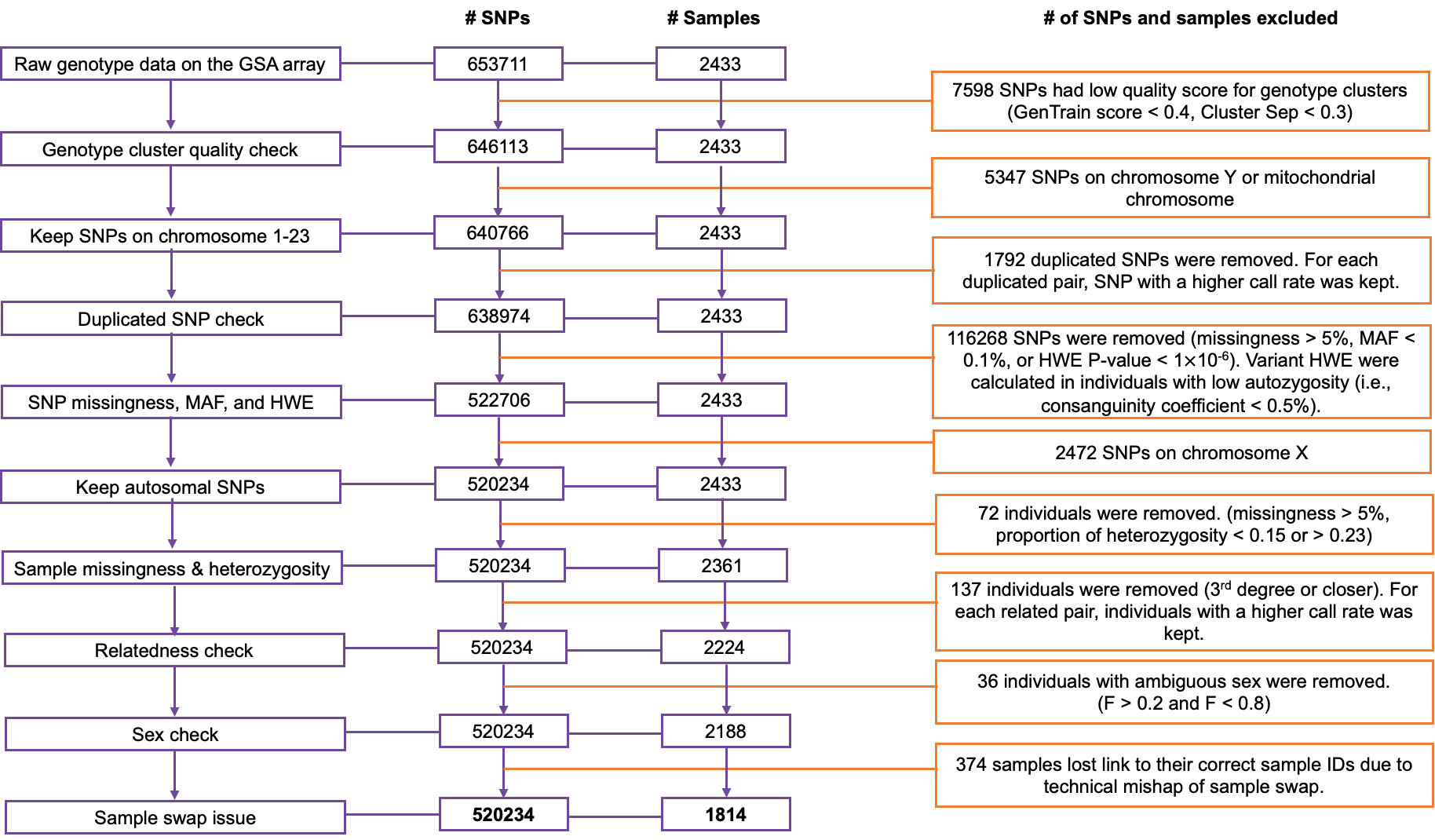

**Figure S2. Quality control (QC) of the Pakistani genotype data before imputation.** After the SNP-level and individual-level QC, a total of 520234 genotyped SNPs for 1814 Pakistani individuals were used for phasing. Abbreviation: GSA, the Illumina Infinium^TM^ Global Screening Array; HWE, Hardy Weinberg Equilibrium; MAF, minor allele frequency; SNP, single nucleotide polymorphism.

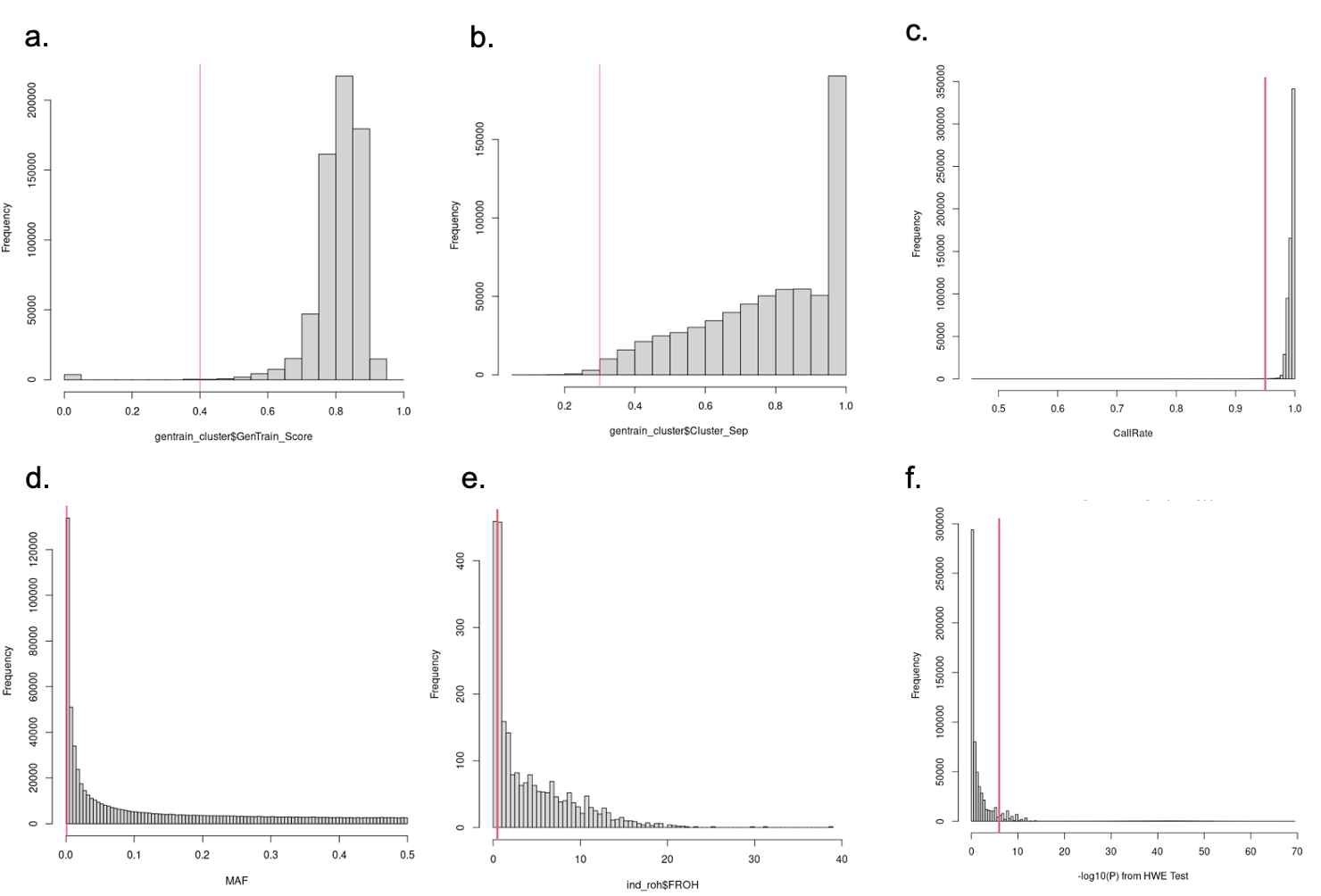

**Figure S3. SNP-level quality control.** (a) SNPs with Gentrain score < 0.4 were removed, (b) SNPs with cluster separation score < 0.3 were removed, (c) SNPs with call rate < 95% were removed, (d) SNPs with MAF < 0.1% were removed, (e) distribution of consanguinity coefficient (F_ROH_) of the Pakistani individuals. A total of 459 out of 2433 Pakistani individuals (19%) have low autozygosity (F_ROH_ < 0.5%). (f) Variants with Hardy Weinberg Equilibrium -log10(p-value) > 6 were identified in the 459 individuals with low autozygosity and removed in all individuals. Abbreviation: HWE, Hardy Weinberg Equilibrium; MAF, minor allele frequency; ROH, runs of homozygosity; SNP, single nucleotide polymorphism.

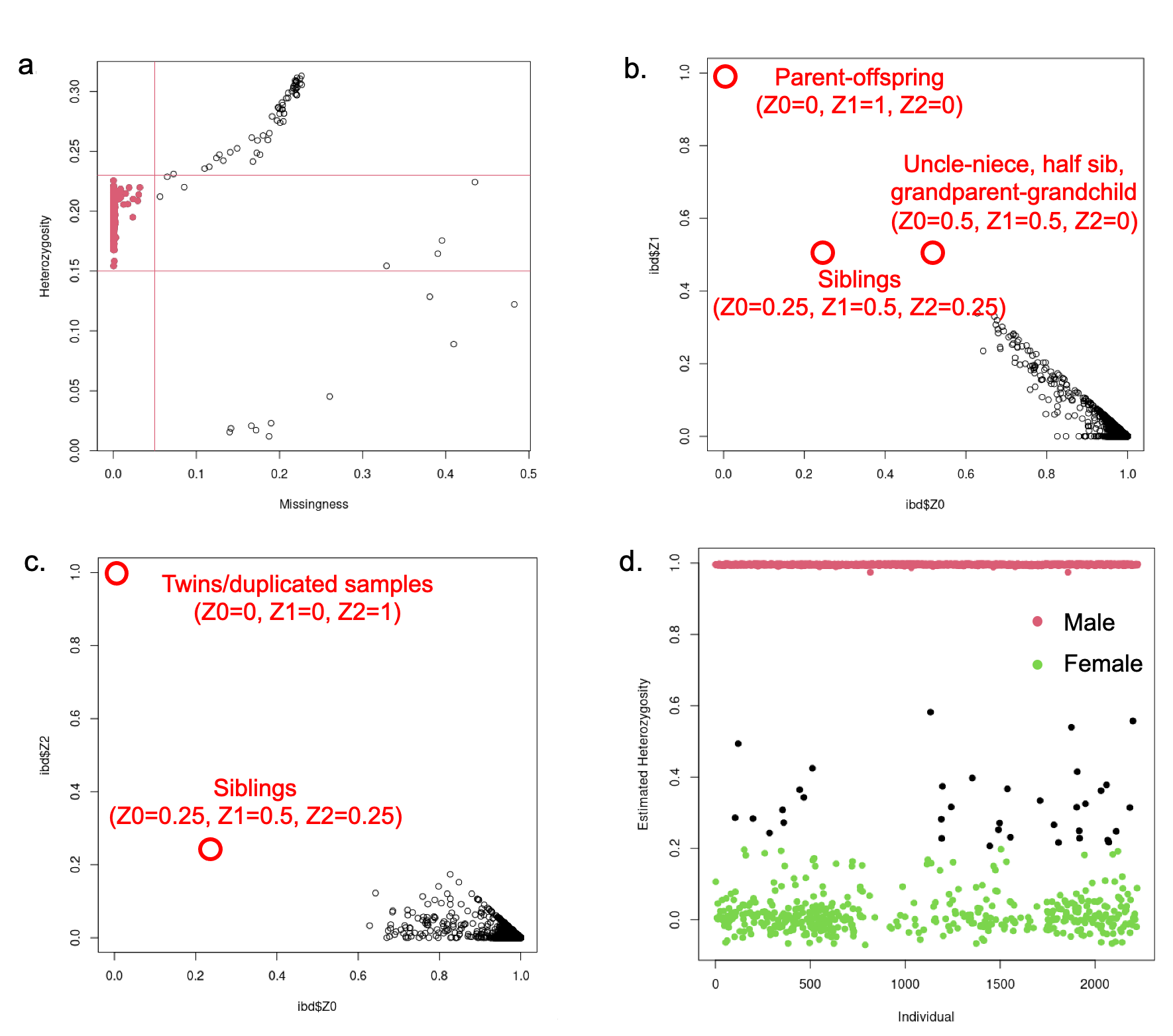

**Figure S4. Individual-level quality control.** (a) A total of 72 Individuals with missingness > 5% and/or heterozygosity proportion outside the range of 0.15 and 0.23 were excluded. (b) Identity-by-descent (IBD) Z0 and Z1 after removing 137 related individuals. Individuals of 3^rd^ degree relatives or closer were removed successfully, indicated by no sample data point on the upper left quadrant. Z0 indicates the probability of not sharing any IBD allele at a locus, whereas Z1 indicates the probability of sharing 1 IBD allele at a locus. For example, in the parent-offspring relationship, the probability of one parent and one offspring sharing one allele at a locus is 100%. (c) Identity-by-descent Z0 and Z2 after removing 137 related individuals. The distribution of data points suggests no pairs of relatives is included now. Z2 means the probability of sharing 2 IBD alleles at a locus. For example, in the case of twins or duplicated samples, the probability of them sharing the same 2 alleles at one locus is 100%. (d) Individuals with ambiguous sex in between heterozygosity F of 0.2 and 0.8 (black dots) were removed.

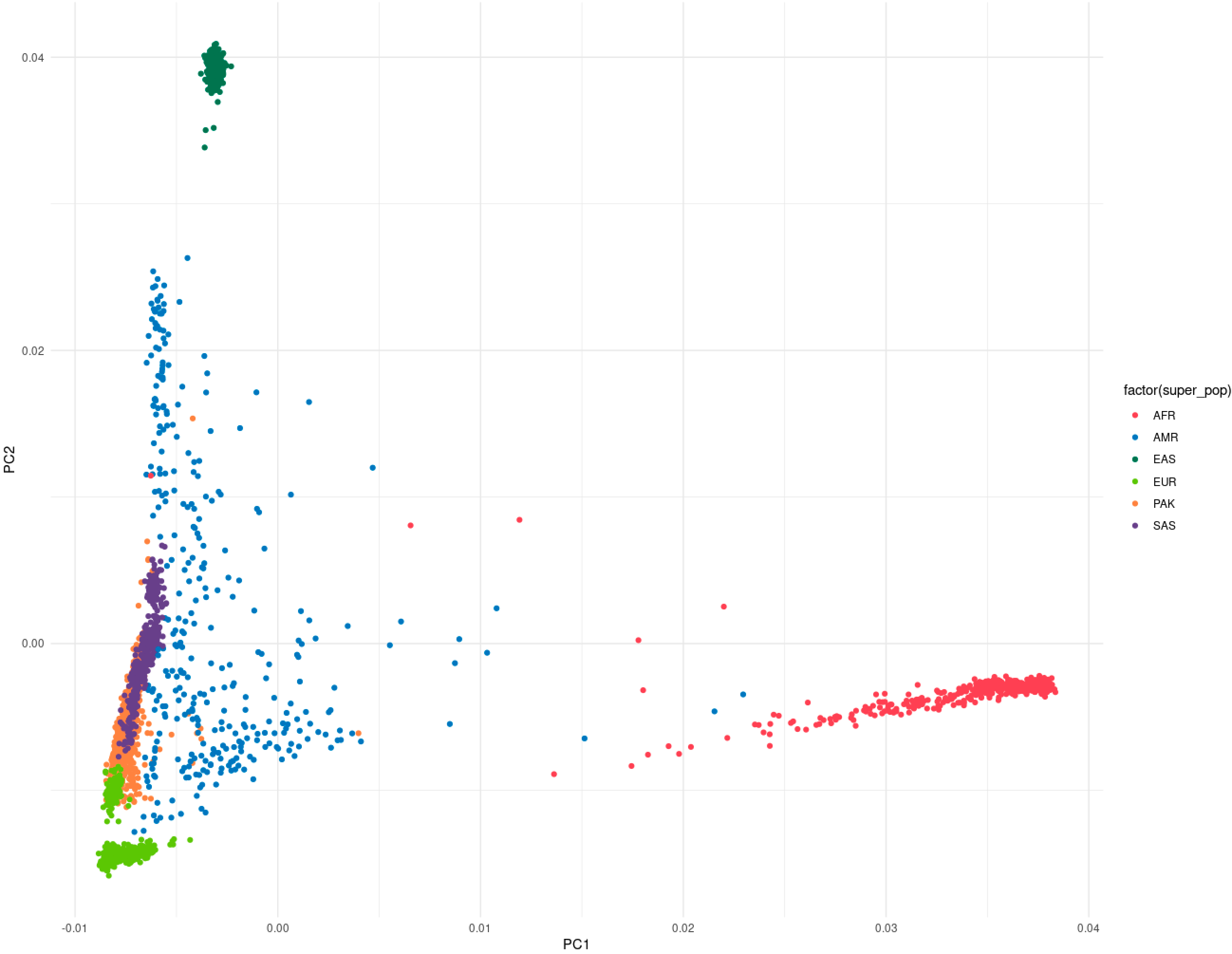

**Figure S5. Pakistani individuals in our study cluster closely with the South Asian population in the 1000 Genomes Project**. This figure plots the first two principal components (PC) based on the genetic data of our samples (PAK) and individuals included in the 1000 Genomes Project phase 3 data (x axis: PC1, y axis: PC2). The Pakistani samples included in this study cluster closely with individuals of South Asian ancestry in the 1000 Genomes Project. The 5 super populations included in the 1000 Genomes Project are African (AFR), admixed American (AMR), East Asian (EAS), European (EUR), and South Asian (SAS).

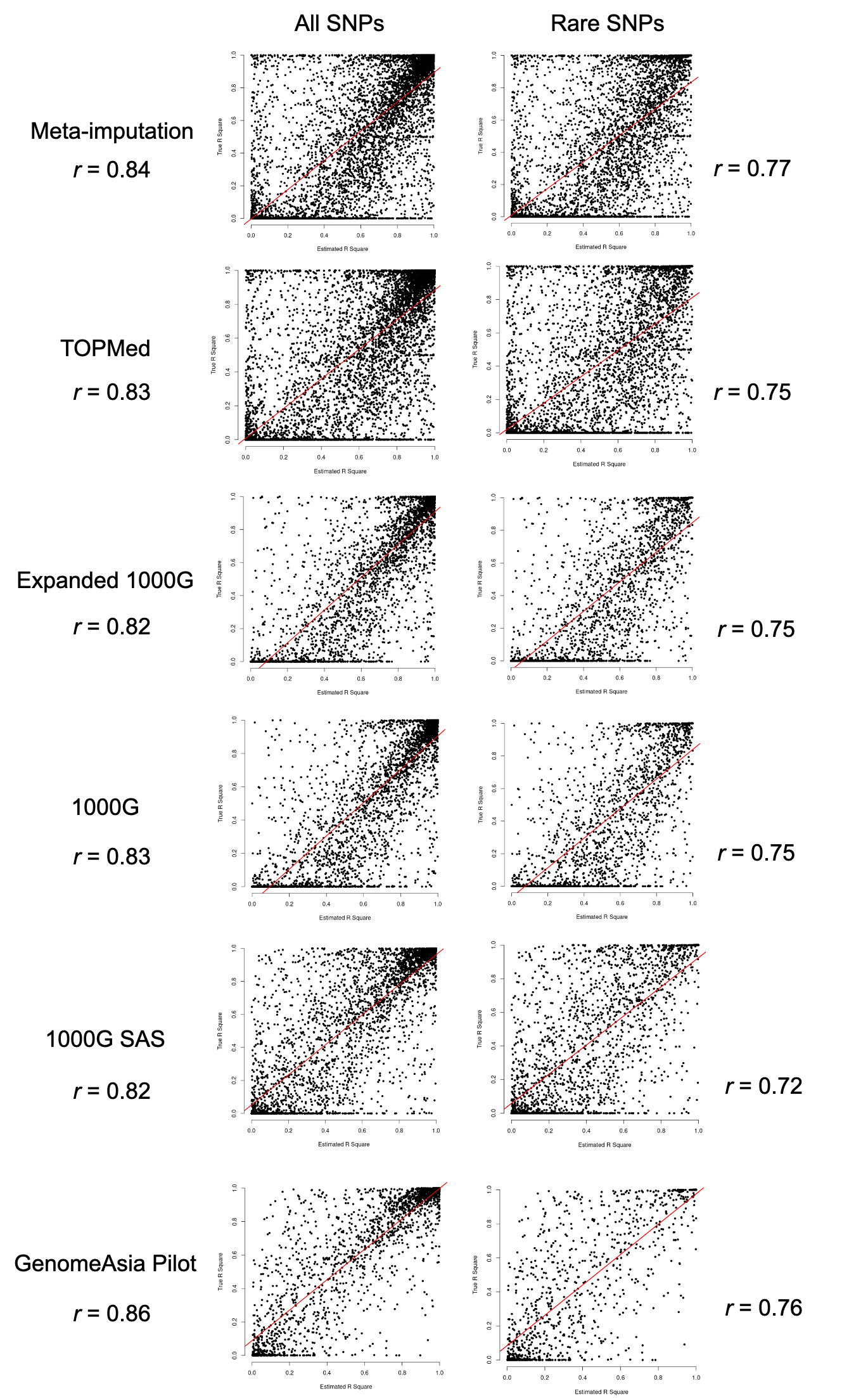

**Figure S6. Correlation between estimated R^2^ and true R^2^.** The left column shows the correlation between $R_{Est}^{2}$ and $R_{True}^{2}$ for each of the SNPs that overlap with sequenced SNPs. The correlation between $R_{Est}^{2}$ and $R_{True}^{2}$ is examined for each of the imputation panels. Overall, $R_{Est}^{2}$ seems to be a good proxy for $R_{True}^{2}$ (*r* > 0.8). The right column shows the correlation between $R_{Est}^{2}$ and $R_{True}^{2}$ for each of the rare SNPs (MAF < 0.01) that overlap with sequenced SNPs. The $R_{Est}^{2}$ is a less accurate proxy for $R_{True}^{2}$ for rare SNPs.

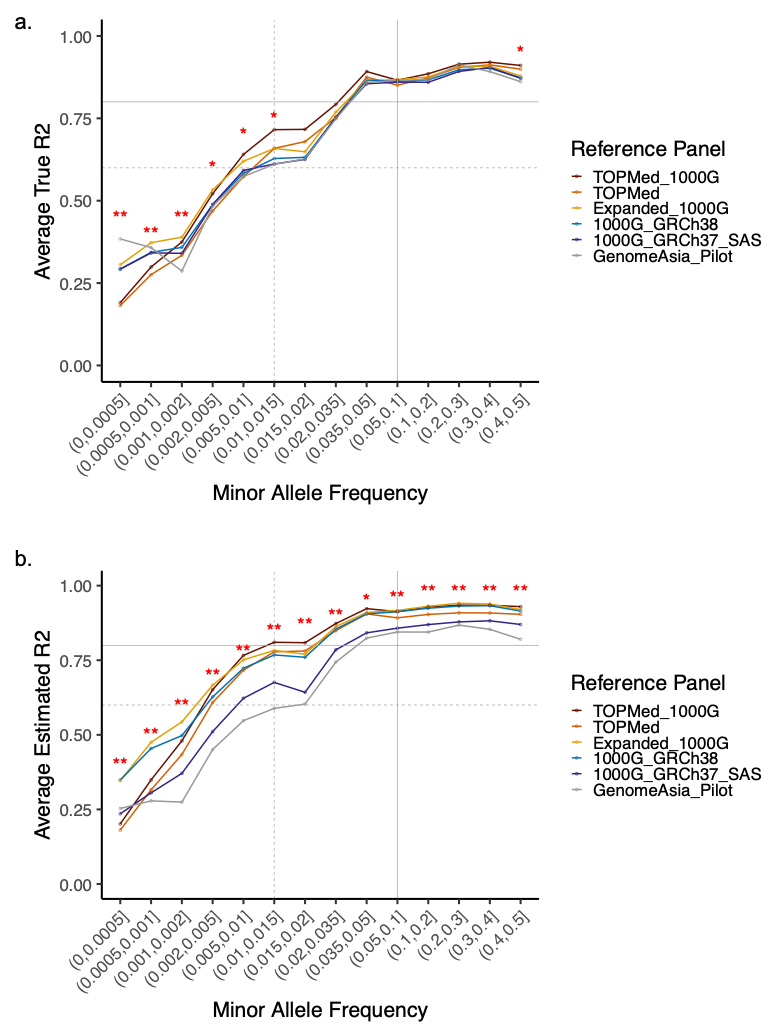

**Figure S7. Comparison of average true R^2^ and average estimated R^2^ across different MAF bins in SNPs that were both sequenced and imputed.** (a) $R_{True}^{2}$ across MAF bins, this is also included as Figure 1a in the main text and is included here for comparison with the $R_{Est}^{2}$ plot. (b) $R_{Est}^{2}$ across MAF bins. The increasing trend is similar for the $R_{True}^{2}$ and $R_{Est}^{2}$ for these SNPs, suggesting that $R_{Est}^{2}$ is a good approximation of $R_{True}^{2}$ in situations where there is no sequencing data to compare against. Overall, the best and worst performing imputation panels are consistent regardless of the imputation metrics used ($R_{True}^{2}$ or $R_{Est}^{2}$). The single asterisk * indicates that there is a nominally significant difference in the average R^2^ across different imputation panels (Kruskal-Wallis rank sum test P < 0.05) at a specific MAF bin, whereas ** denotes that there is a Bonferroni-corrected significant difference (P < 0.003, correcting for the number of pairwise panels tested). The horizontal grey lines indicate R^2^ of 0.6 (dashed line, indicating fairly high imputation quality) and 0.8 (solid line, indicating high imputation quality). The vertical grey lines indicate MAF > 1% (dashed line) and > 5% (solid line). Abbreviation: 1000G, 1000 Genomes; MAF, minor allele frequency; R^2^, squared correlation; SAS, South Asian ancestry; SNP, single nucleotide polymorphism.

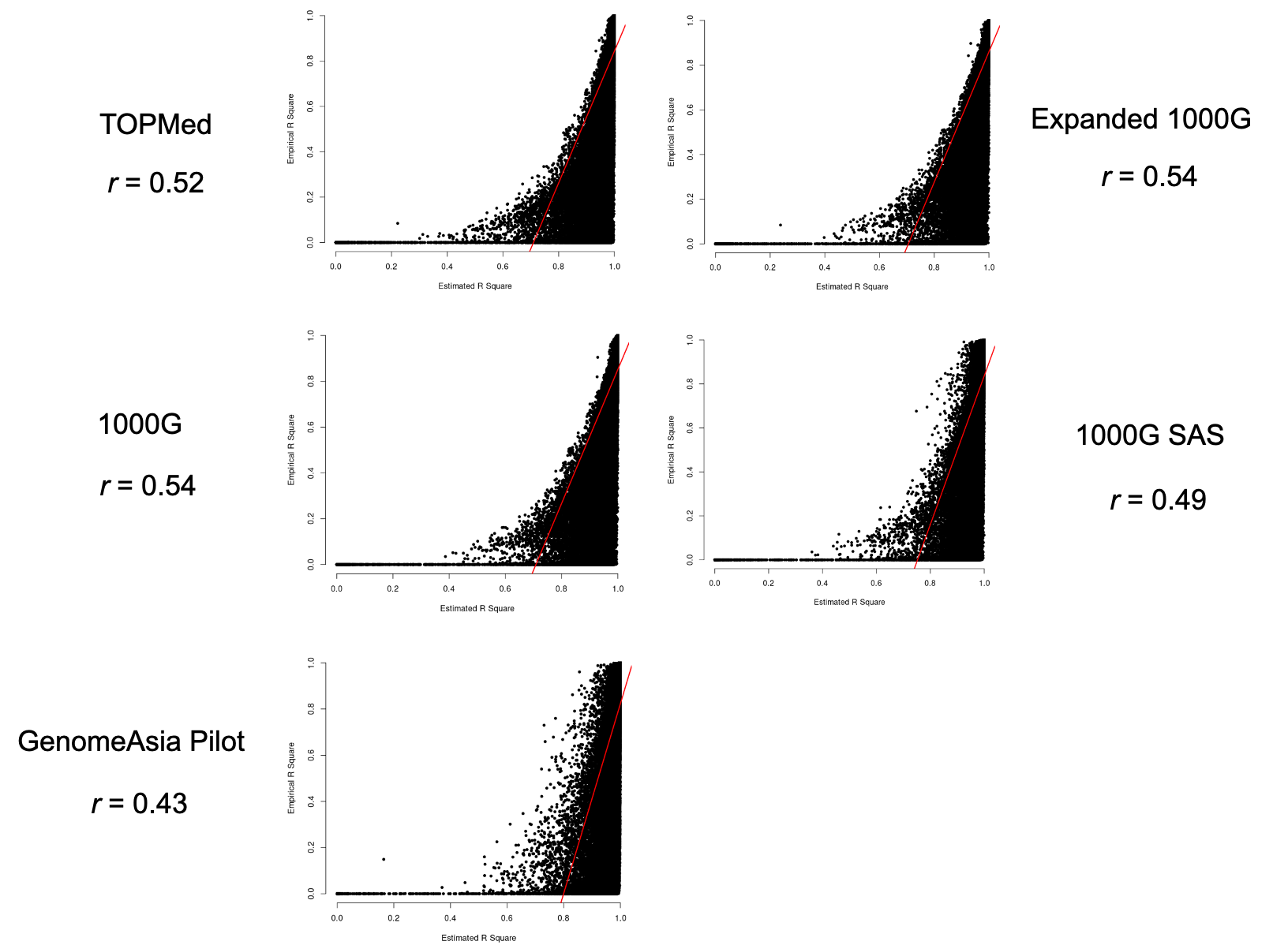

**Figure S8. Correlation between empirical R^2^ and estimated R^2^.** Each figure shows the correlation between $R_{Emp}^{2}$ and $R_{Est}^{2}$ for each of the genotyped SNPs. $R_{Emp}^{2}$ for each SNP is the squared correlation between the directly measured genotype by the GSA array and the imputed genotype by each imputation panel. The $R_{Est}^{2}$ for each SNP is calculated based on posterior allele probabilities. The correlation between $R_{Emp}^{2}$ and $R_{Est}^{2}$ is examined for each imputation panel, except meta-imputation, which does not produce $R_{Emp}^{2}$ values. Overall, $R_{Emp}^{2}$ does not correlate well with $R_{Est}^{2}$ (*r* ranges from 0.43 to 0.54 for different imputation panels). Abbreviation: 1000G, 1000 Genomes; GSA, the Illumina Infinium^TM^ Global Screening Array; SAS, South Asian ancestry; SNP, single nucleotide polymorphism.

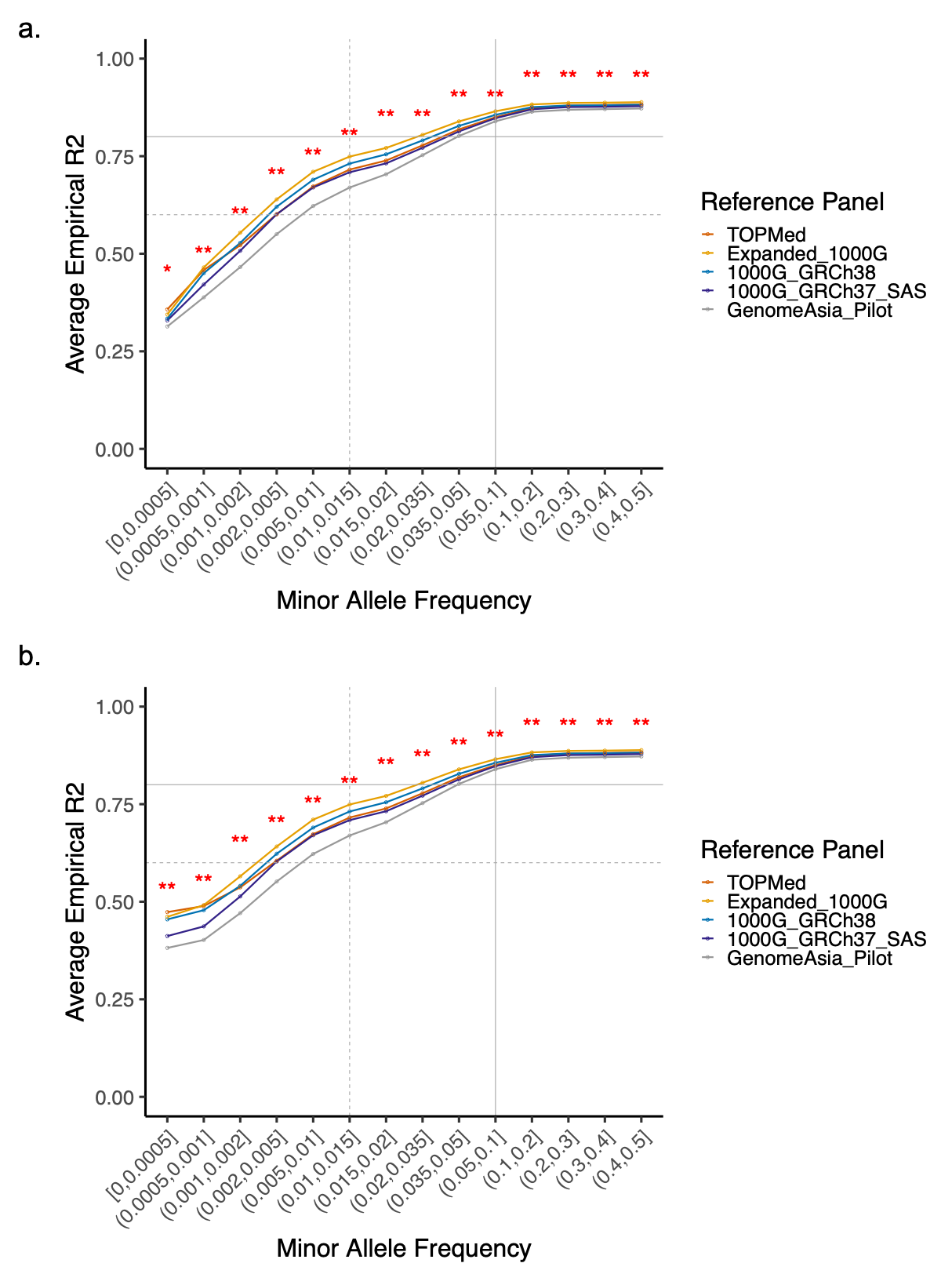

**Figure S9. Empirical R^2^ increases with MAF for each imputation panel.** (a) $R_{Emp}^{2}$ of directly genotyped SNPs that also have imputed dosages. (b) $R_{Emp}^{2}$ in directly genotyped SNPs that have well-imputed dosages with estimated R^2^ $\geq$ 0.8. When limited to SNPs with high $R_{Est}^{2}$, unlike $R_{True}^{2}$, the average $R_{Emp}^{2}$ has only minor improvement, indicating the poor correlation between $R_{Est}^{2}$ and $R_{Emp}^{2}$. The single asterisk * indicates that there is a nominally significant difference in the average $R_{Emp}^{2}$ across different imputation panels (Kruskal-Wallis rank sum test P < 0.05) at a specific MAF bin, whereas ** denotes that there is a Bonferroni-corrected significant difference (P < 0.003, correcting for the number of pairwise panels tested). The horizontal grey lines indicate $R_{Emp}^{2}$ of 0.6 (dashed line, indicating fairly high imputation quality) and 0.8 (solid line, indicating high imputation quality). The vertical grey lines indicate MAF > 1% (dashed line) and > 5% (solid line). Abbreviation: 1000G, 1000 Genomes; MAF, minor allele frequency; R^2^, squared correlation; SAS, South Asian ancestry; SNP, single nucleotide polymorphism.

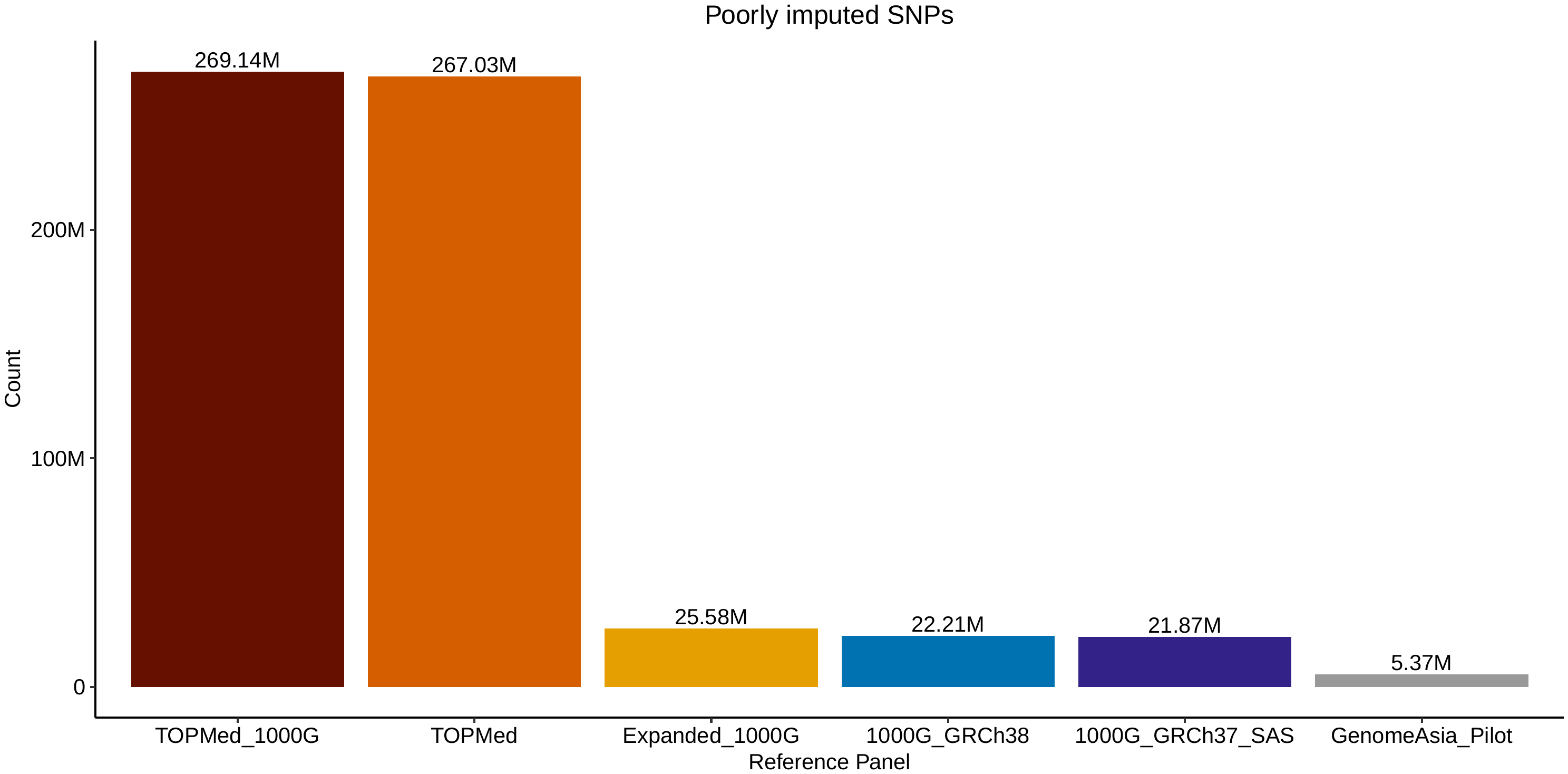

**Figure S10. Number of imputed ultra-rare variants with MAF** $\boldsymbol{\leq}$ **0.05% and estimated R^2^ < 0.8 across imputation panels.** Abbreviation: 1000G, 1000 Genomes; MAF, minor allele frequency; SAS, South Asian ancestry; SNP, single nucleotide polymorphism.

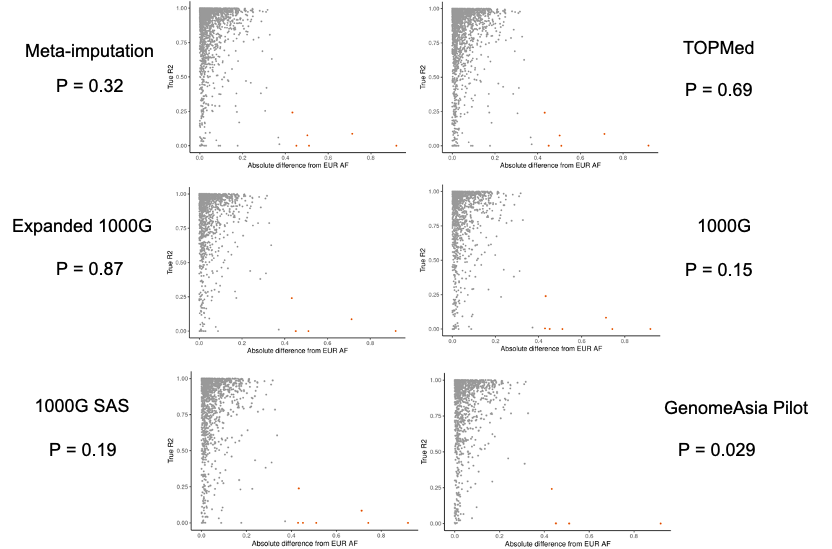

**Figure S11. The deviation from EUR AF overall does not impair imputation quality of common SNPs, measured by true R^2^, in Pakistani individuals.** The common SNPs are defined as SNPs with MAF $\geq$ 1% measured by targeted sequencing in the Pakistani individuals. The x axis is the absolute difference in allele frequency between Pakistani individuals (measured by targeted sequencing) and European ancestry individuals based on 1000 Genomes data. The y axis is the imputation quality of the SNPs, measured by $R_{True}^{2}$. Unlike our hypothesis that SNPs deviated from EUR AF will suffer in imputation accuracy by current imputation panels, for common SNPs, we observed that after Bonferroni correction, none of the associations was significant (P_Bonferroni_ = 0.05/6 panels = 0.0083), such that there is no association between deviation from EUR AF and imputation quality. Outlier SNPs highlighted in red (ranging from 4 to 7 SNPs across different imputation panels) have low imputation quality scores due to the discrepancy in AF based on the imputed genotype versus AF based on the sequenced genotype in the Pakistani individuals, rather than because of the deviation from EUR AF (more details on the imputed and sequenced AF of these SNPs in the Pakistani individuals are listed in Table S5). Abbreviation: 1000G, 1000 Genomes; AF, allele frequency; EUR, European ancestry; MAF, minor allele frequency; SAS, South Asian ancestry; SNP, single nucleotide polymorphism.

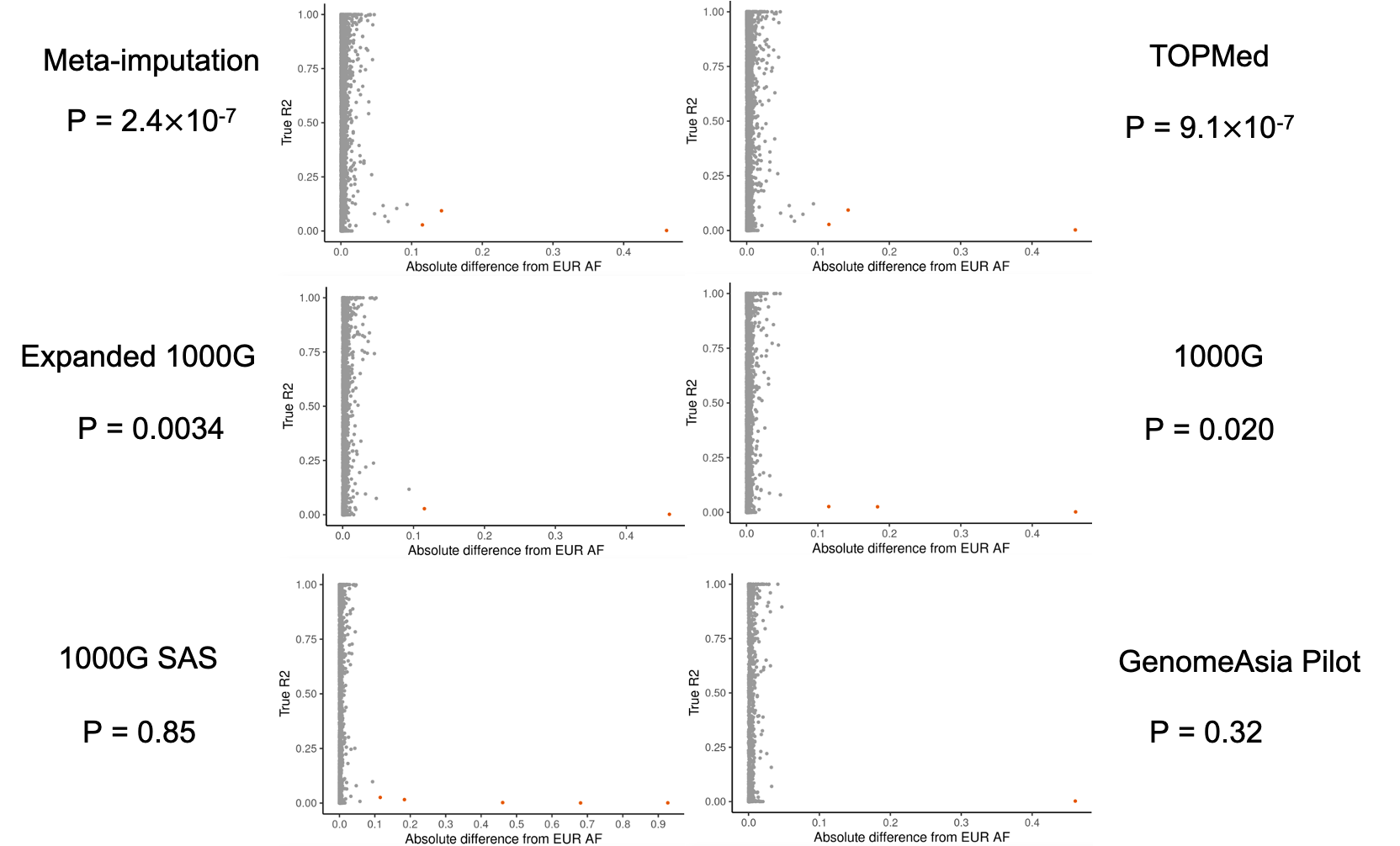

**Figure S12. The deviation from EUR AF overall does not impair imputation quality of rare SNPs, measured by true R^2^, in Pakistani individuals.** The rare SNPs are defined as SNPs with MAF less than 1% measured by targeted sequencing in the Pakistani individuals. The x axis is the absolute difference in allele frequency between Pakistani individuals (measured by targeted sequencing) and European ancestry individuals based on 1000 Genomes data. The y axis is the imputation quality of the SNPs, measured by $R_{True}^{2}$. Unlike our hypothesis that SNPs deviated from EUR AF will suffer in imputation accuracy by current imputation panels, for rare SNPs, we observed either a *positive* association such that SNPs with greater deviation from EUR AF have greater $R_{True}^{2}$ values, or a *null* association where there is no association between deviation from EUR AF and imputation quality. Outlier SNPs highlighted in red (ranging from 1 to 5 SNPs across different imputation panels) have low imputation quality scores due to the discrepancy in AF based on the imputed genotype versus AF based on the sequenced genotype in the Pakistani individuals, rather than because of the deviation from EUR AF (more details on the imputed and sequenced AF of these SNPs in the Pakistani individuals are listed in Table S5). Abbreviation: 1000G, 1000 Genomes; AF, allele frequency; EUR, European ancestry; MAF, minor allele frequency; SAS, South Asian ancestry; SNP, single nucleotide polymorphism.

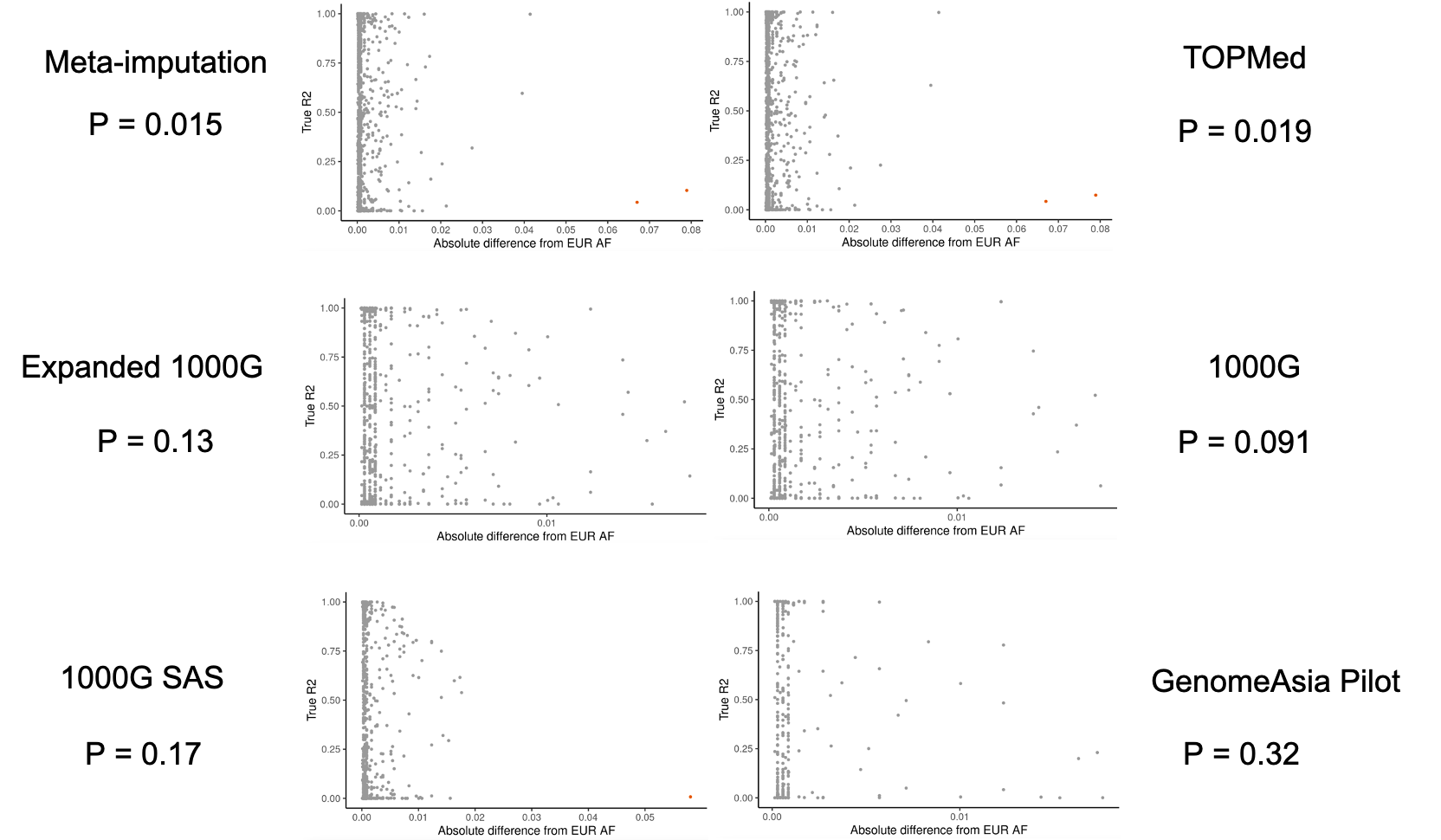

**Figure S13. The deviation from EUR AF overall does not impair imputation quality of ultra-rare SNPs, measured by true R^2^, in Pakistani individuals.** The ultra-rare SNPs are defined as SNPs with MAF less than 0.1% measured by targeted sequencing in the Pakistani individuals. The x axis is the absolute difference in allele frequency between Pakistani individuals (measured by targeted sequencing) and European ancestry individuals based on 1000 Genomes data. The y axis is the imputation quality of the SNPs, measured by $R_{True}^{2}$. Unlike our hypothesis that SNPs deviated from EUR AF will suffer in imputation accuracy by current imputation panels, for ultra-rare SNPs, we observed that after Bonferroni correction, none of the associations was significant (P_Bonferroni_ = 0.05/6 panels = 0.0083), such that there is no association between deviation from EUR AF and imputation quality. Outlier SNPs highlighted in red (ranging from 0 to 2 SNPs across different imputation panels) have low imputation quality scores due to the discrepancy in AF based on the imputed genotype versus AF based on the sequenced genotype in the Pakistani individuals, rather than because of the deviation from EUR AF (more details on the imputed and sequenced AF of these SNPs in the Pakistani individuals are listed in Table S5). Abbreviation: 1000G, 1000 Genomes; AF, allele frequency; EUR, European ancestry; MAF, minor allele frequency; SAS, South Asian ancestry; SNP, single nucleotide polymorphism.

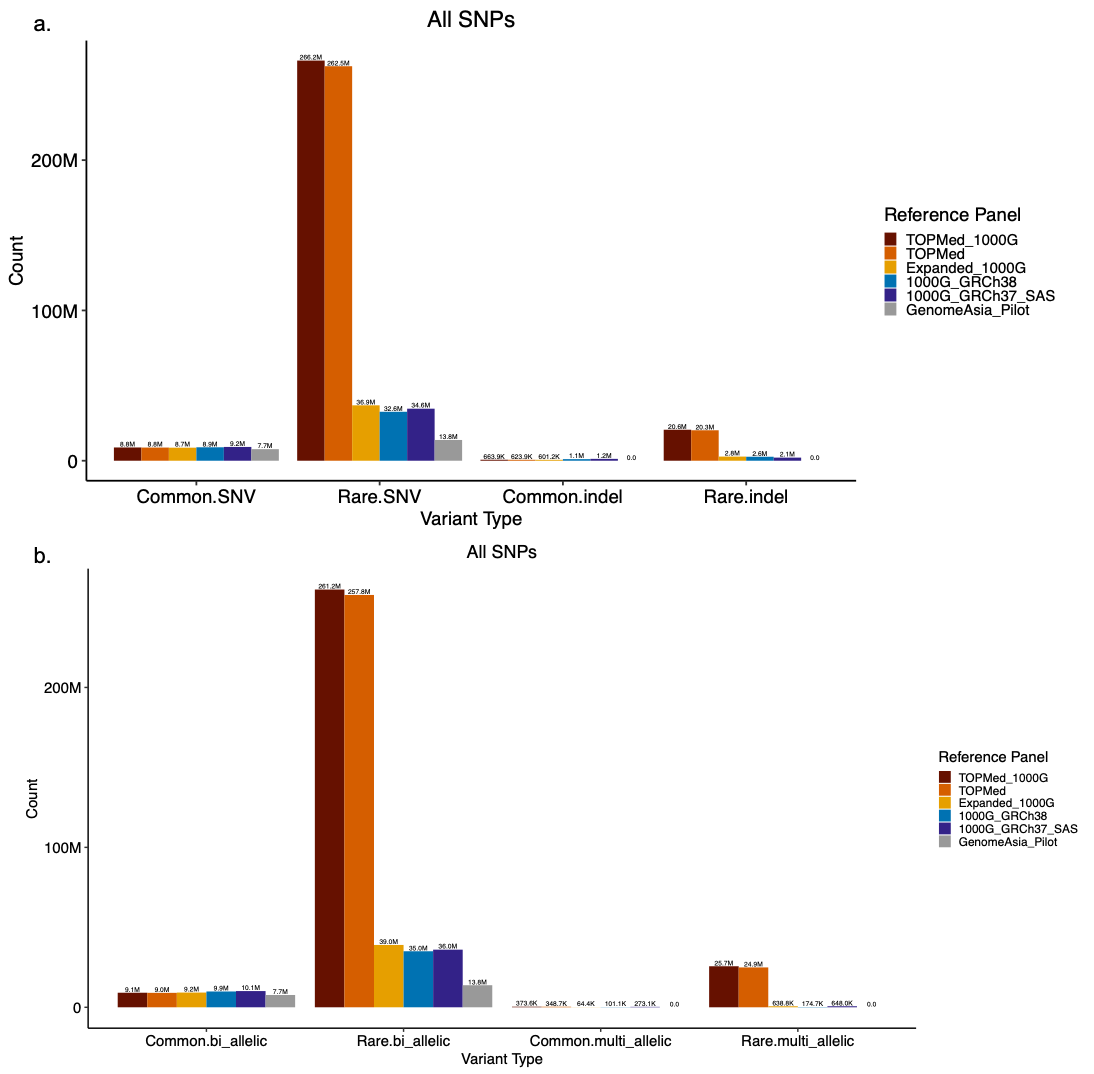

**Figure S14. Genome-wide imputed SNP counts by different SNP types in the Pakistani individuals.** a) The number of imputed SNPs genome-wide by common/rare and SNV/indel status; b) The number of imputed SNPs genome-wide by common/rare and bi-allelic/multi-allelic status. Abbreviation: 1000G, 1000 Genomes; indel, insertion-deletion; SAS, South Asian ancestry; SNP, single nucleotide polymorphism; SNV, single nucleotide variant.

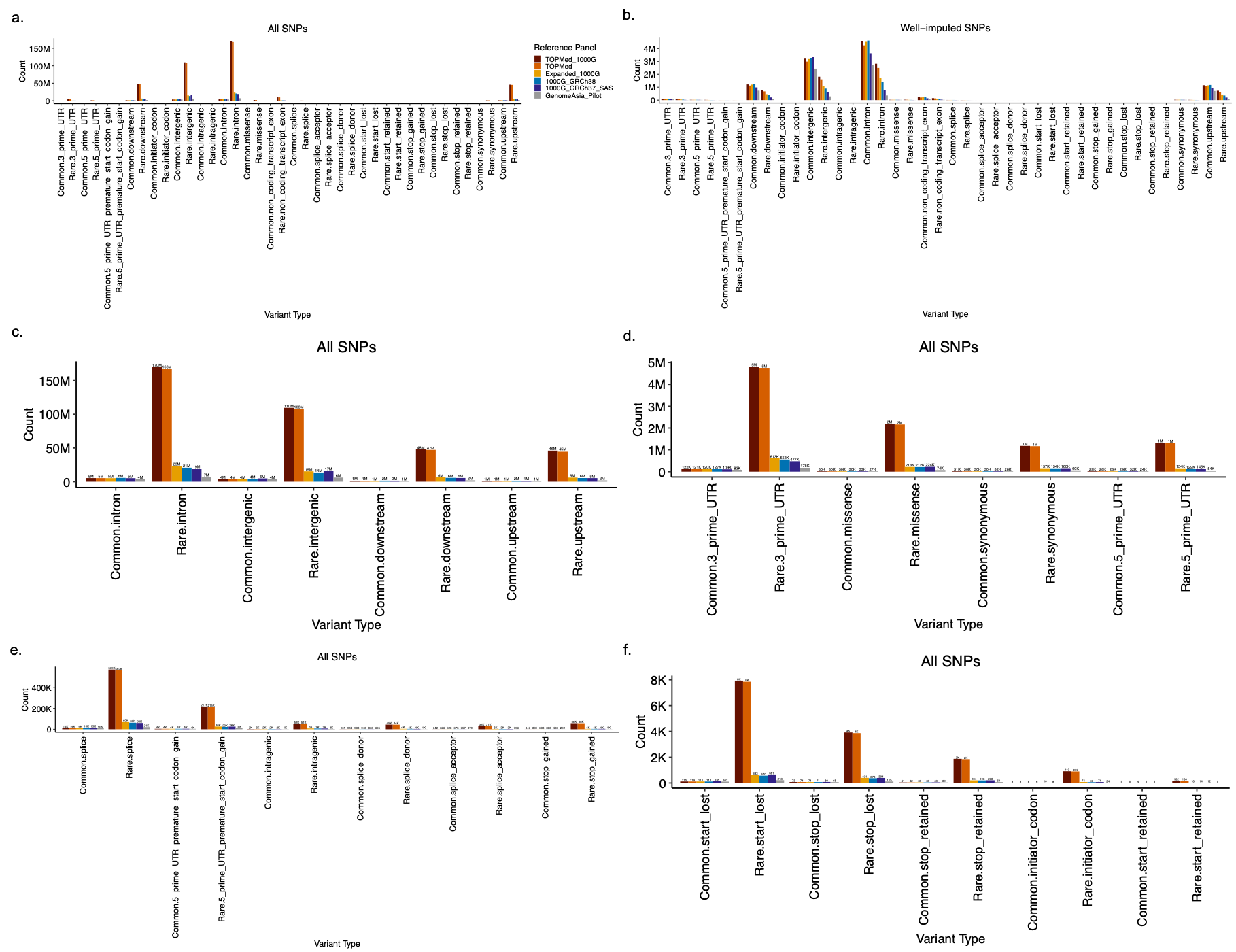

**Figure S15. Genome-wide imputed SNP counts by different SNP types via functional annotation in the Pakistani individuals.** a) The number of imputed SNPs genome-wide by common/rare status and different SNP types; b) The number of well-imputed SNPs (defined as estimated R^2^ $\geq$ 0.8) genome-wide by common/rare status and different SNP types; c-f) The number of imputed SNPs genome-wide by common/rare status and SNP types; c) includes 4 SNP types with the highest SNP counts (i.e., intron, intergenic, downstream, upstream); d) includes 5 SNP types with following highest SNP counts (i.e., non-coding transcript exon, 3’ UTR, missense, synonymous, 5’ UTR); e) includes 6 SNP types with low SNP counts (i.e., splice region, 5’ UTR premature start codon gain, intragenic, splice donor, splice acceptor, stop gained); f) includes 5 SNP types with ultra-low SNP counts (i.e., start lost, stop lost, stop retained, initiator codon, start retained). Abbreviation: 1000G, 1000 Genomes; SAS, South Asian ancestry; SNP, single nucleotide polymorphism; UTR, untranslated region.
